# Global diversity of policy, coverage, and demand of COVID-19 vaccines: a descriptive study

**DOI:** 10.1101/2021.10.25.21265504

**Authors:** Zhiyuan Chen, Wen Zheng, Qianhui Wu, Xinghui Chen, Cheng Peng, Yuyang Tian, Ruijia Sun, Minghan Wang, Xiaoyu Zhou, Zeyao Zhao, Guangjie Zhong, Xuemei Yan, Nuolan Liu, Feiran Hao, Sihong Zhao, Tingyu Zhuang, Juan Yang, Andrew S. Azman, Hongjie Yu

## Abstract

**Background:** Hundreds of millions of doses of COVID-19 vaccines have been administered globally, but progress in vaccination varies considerably between countries. We aim to provide an overall picture of COVID-19 vaccination campaigns, including policy, coverage, and demand of COVID-19 vaccines.

**Methods:** We conducted a descriptive study of vaccination policy and doses administered data obtained from multiple public sources as of 23 October 2021. We used these data to develop coverage indicators and explore associations of vaccine coverage with socioeconomic and healthcare-related factors. We estimated vaccine demand as numbers of doses required to complete vaccination of countries’ target populations according to their national immunization program policies.

**Findings:** Use of both mRNA and adenovirus vectored vaccines was the most commonly used COVID-19 vaccines formulary in high-income countries, while adenovirus vectored vaccines were the most widely used vaccines worldwide (176 countries). Almost all countries (98.3%, 173/176) have authorized vaccines for the general public, with 53.4% (94/176) targeting individuals over 12 years and 33.0% (58/176) targeting those ≥18 years. Forty-one and sixty-seven countries have started additional-dose and booster-dose vaccination programs, respectively. Globally, there have been 116.5 doses administered per 100 target population, although with marked inter-region and inter-country heterogeneity. Completed vaccination series coverage ranged from 0% to more than 95.0% of country target populations, and numbers of doses administered ranged from 0 to 239.6 per 100 target population. Doses administered per 100 total population correlated with healthcare access and quality index (R^2^ = 0.58), socio-demographic index (R^2^ = 0.56), and GDP per capita (R^2^ = 0.65). At least 5.54 billion doses will be required to complete interim vaccination programs – 4.65 billion for primary immunization and 0.89 billion for additional/booster programs. Globally, 0.84 and 0.96 dose per individual in the target population are needed for primary immunization and additional/booster programs, respectively.

**Interpretation:** There is wide country-level disparity and inequity in COVID-19 vaccines rollout, suggesting large gaps in immunity, especially in low-income countries.

**Funding:** Key Program of the National Natural Science Foundation of China, the US National Institutes of Health.

**Research in context:** *Evidence before this study:* We searched PubMed for articles in any language published up to October 21, 2021, using the following search terms: (“COVID-19” OR “SARS-CoV-2”) AND (“vaccination” OR “vaccine”) AND (“inequalit*” OR “inequity” OR “disparit*” OR “heterogeneity”). We also searched for dashboards associated with vaccine rollout from public websites. We identified several studies on tracking global inequalities of vaccine access, one of which constructed a COVID-19 vaccine dashboard (Our World in Data), and another that explored disparities in COVID-19 vaccination among different-income countries. However, we found no studies that depict global COVID-19 vaccination policies country-by-country and estimate demand for vaccine necessary to completely vaccinate countries’ designated target populations.

*Added value of this study:* To our knowledge, our study provides the most recent picture of COVID-19 vaccination campaigns, focusing on global vaccination policy and target-population demand. We found a diverse portfolio of vaccines in five technical platforms being administered globally, with 173 countries having authorized administration of vaccines to the general public in various age groups. We observed inter-region and inter-country heterogeneity in one-or-more-dose and full-dose coverage; countries with higher socio-demographic or health resource-related levels had higher coverage. We estimated dose-level demand for completing primary immunization programs and additional/booster dose programs separately.

*Implications of all the available evidence:* Worldwide disparity and inequity of vaccine rollout implies that susceptibility among unvaccinated populations in some countries may impede or reverse pandemic control, especially in face of the emergence of variants and the dilemma of waning antibodies. Our findings suggest that global-level responses to the pandemic - financially, politically, and technically - are needed to overcome complex challenges that lie ahead.

## Introduction

The Coronavirus Disease 2019 (COVID-19) pandemic is still raging globally and has had an unprecedented impact on societies and economies^1^. Several non-pharmaceutical interventions (NPIs) have been effective to reduce virus transmission^2,3^, but it is unrealistic to continue many of these NPIs for long-term maintenance. Rapid and successful development of efficacious and effective vaccines against COVID-19 is making it possible to manage the pandemic^4^.

General public health use of COVID-19 vaccines started in December 2020 and has accelerated globally at an unprecedented rate. However, inequitable country-level access to vaccines may unbalance global immunity and will likely impede global reopening. People may not get vaccinated for a variety of reasons, including supply-and-demand challenges, country purchase capacity, not being a member of a country-specific vaccine target population, and personal hesitancy to receive vaccine^5-8^. Most high-income countries have purchased vaccine directly using advanced purchase agreements with vaccine developers and manufacturers^5^. Low-income countries were less able to purchase/receive vaccines as needed, although global efforts like COVID-19 Vaccine Global Access (COVAX)^9^, are starting to increase access.

Disparities in COVID-19 vaccination progress have been observed between and within countries. Several dashboards use official data sources to track vaccination progress, but there has been insufficient focus on variation in countries’ vaccination policies^1,10^. One study explored the correlation between GDP per capita and vaccine coverage in 138 countries and showed disparities in vaccine rollout among countries with different income levels^11^. Country-level spatiotemporal disparities in vaccination were also seen^12,13^. An overview of vaccination campaigns across countries, especially of vaccination policy and demand, has not been fully explored.

In this study, we aimed to describe the global landscape of COVID-19 vaccination policy by authorised vaccines, primary/booster vaccination, and target population. We constructed metrics for vaccine coverage to explore the extent and variation in global and regional vaccination progress and estimate demand for vaccine.

## Methods

We constructed datasets for vaccination policy and doses administered as of 23 October 2021 that contain country-specific metrics. We used these metrics to develop three coverage indicators and estimated demand for vaccine doses. Detailed descriptions of data collection methods and data sources are provided in the Appendix.

### Metrics

Policy metrics include authorization status of COVID-19 vaccines (approved, conditionally approved, or authorized for emergency use), vaccination schedules, indicated age groups, contraindications, whether vaccines are sold/donated by or received by the country, and whether vaccines are provided at no cost to individual vaccine recipients. Vaccination schedules are defined in terms of the number of required doses and the dosing intervals according to regulatory authorities, and special doses, such as additional doses for immunocompromised individuals and booster doses. Contraindications are medical conditions for which individuals should not be vaccinated, either temporarily for conditions that resolve, or permanently.

The doses-administered dataset included four time-varying, age-specific, platform-specific metrics: number of vaccine doses administered, number of people receiving at least one dose, number of people fully vaccinated according to country schedule, and number of people receiving additional/booster doses. People receiving at least one dose are those who have received the first dose of a multiple-dose vaccine or the dose of a one-dose vaccine. Number of people fully vaccinated indicates those who have completed their primary immunization series according to the vaccination schedule. Additional doses are doses for people with medical conditions requiring doses beyond the primary series to achieve immunity (e.g., moderately to severely immunocompromised people); booster doses are doses given after the primary series to counter waning immunity or protection^14^.

### Target populations

Target populations are eligible individuals who are within approved age groups and without contraindications according to the country’s immunization policy. When country-specific information was unavailable, we used the WHO-recommended ages, in which people 12 years and older are recommended to receive Pfizer-BioNTech vaccine and people 18 years and above are recommended to receive any COVID-19 vaccine^15^. Contraindications vary by country, and may include pregnant women^16^, people with certain underlying conditions (i.e. bleeding disorders and immune suppression)^17^, and/or those previously infected with SARS-CoV-2^1^. Details are in Table S2 and Table S6. We estimated sizes of target populations for additional and booster doses according to country-specific policy (Table S3), using Clark’s method to eliminate the effect of multimorbidity^17^. Methods for estimating target population sizes are detailed in the Appendix.

### Vaccine coverage

We constructed vaccine coverage indicators among total populations and target populations; indicators are full vaccination coverage, share of receipt of at least one dose, and doses per 100 people. Because not all countries report vaccine doses administered on a daily basis, we used linear extrapolation between reported data points. We estimated vaccine coverage up to 7 October 2021. For countries that did not report data to 7 October, we used data reported on or after 18 September 2020, or missing values if no data were reported on/after 18 September.

### Factors associated with coverage

To investigate factors related with vaccine coverage, we identified a list of economic, social, health spending, health resources-related factors potentially associated with vaccine coverage. Due to multicollinearity, we used socio-demographic index (SDI), healthcare access and quality (HAQ) Index, and GDP per capita adjusted for purchasing power parity (PPP) in our analyses. SDI is a composite indicator developed by the Institute for Health Metrics and Evaluation (IHME), containing lag-distributed income per capita, average education level, and fertility rate under 25 years; it ranges from 0 to 1 and can be divided into five categories: high, high-middle, middle, low-middle, and low SDI^18^. HAQ is a country-specific index that quantifies the accessibility and quality of personal healthcare; it ranges from 0 to 100^19^. We obtained GDP per capita-PPP for 2019 from the World Bank^20^.

### Demand

We determined demand for vaccine as the number of doses needed to complete vaccination of countries’ target populations according to national immunization program policy; we estimate demand for primary immunization and additional/booster doses separately. We used a simplifying assumption that all COVID-19 vaccines primary series require two doses, as one-dose (Janssen COVID-19 Vaccine, Convidecia) and three-dose (Zifivax, Abdala) vaccines are thus far a small (though unknown) proportion of total doses administered. We estimated additional and booster doses needed based on sizes of certain populations allowed by regulatory agencies; we assumed one additional/booster dose per person recommended or indicated. We calculated total current demand as the sum of doses required minus doses administered by 7 October 2021, stratified by primary and additional/booster dose demand.

### Data sources

In priority order, we obtained data from government websites, health department websites, official media, vaccine manufacturers’ websites, authoritative media, and local media. We also use data from public databases that systematically collect and cross-check such information, such as Gavi-COVAX^21^, COVID-19 Vaccine Market Dashboard^22^, and Our World in Data^23^. Whether countries sell/donate or receive vaccine was obtained from COVAX data^21^.

We collected raw data through a combination of manual and automated means. For countries that release data on a regular basis through official sources^24,25^, we collected information manually on a weekly basis; for countries that release official data through public online dataset (e.g., GitHub repository)^26^, we accessed data with a compiled R Script. Countries were included in our doses administered datasets if they reported at least two data points. We verified data with double-entry.

We derived 2020 population estimates from UN World Population Prospects^27^. Among 194 Member States of the World Health Organization (WHO), age-specific UN population proportions were available for 183 countries. Population estimates for the 11 remaining countries (Andorra, Cook Islands, Dominica, Saint Kitts and Nevis, Monaco, Marshall Islands, Niue, Nauru, Palau, San Marino, and Tuvalu) were compiled from WorldPop datasets^28^.

### Statistical analysis

Since the obvious non-linear relationship between vaccine coverage and selected independent factors, we used a nonlinear least squares regression model (logistic growth model) to determine the adjusted relation. All statistical analyses and visualizations were done using R (version 4.0.2).

## Results

### COVID-19 vaccination policy

As of 23 October 2021, 192 countries reported the COVID-19 vaccines they use, and 176 countries reported their target populations. 173 countries have extended vaccination to the general public, and three countries (Central African Republic, Burundi, and Gambia) target only specific groups such as people at risk of severe illness, elderly people, and healthcare workers (Table S2).

Among the five technological vaccine platforms with vaccines approved in one or more country, adenovirus vectored vaccines were the most widely used (176 countries), followed by mRNA vaccines (139 countries), inactivated vaccines (109 countries), protein subunit vaccines (10 countries), and conjugate vaccines (3 countries) (Fig 1B-D). From a regional perspective, high-income countries in Europe and the Americas most often used both mRNA and adenovirus vectored vaccines; other countries in the Americas also used inactivated vaccines (Fig 1A). In Africa, vaccination was primarily with an adenovirus vectored vaccine or a combination of inactivated and adenovirus vectored vaccines; 19 African countries used mRNA vaccines (Fig 1A). Among 30 countries (18 high-income) that use vaccines made by two or more platforms, more than half of the vaccines administered in 76.7% (23) of these countries were mRNA vaccines, mainly Pfizer-BioNTech vaccine (Fig S1-2).

**Figure 1.**
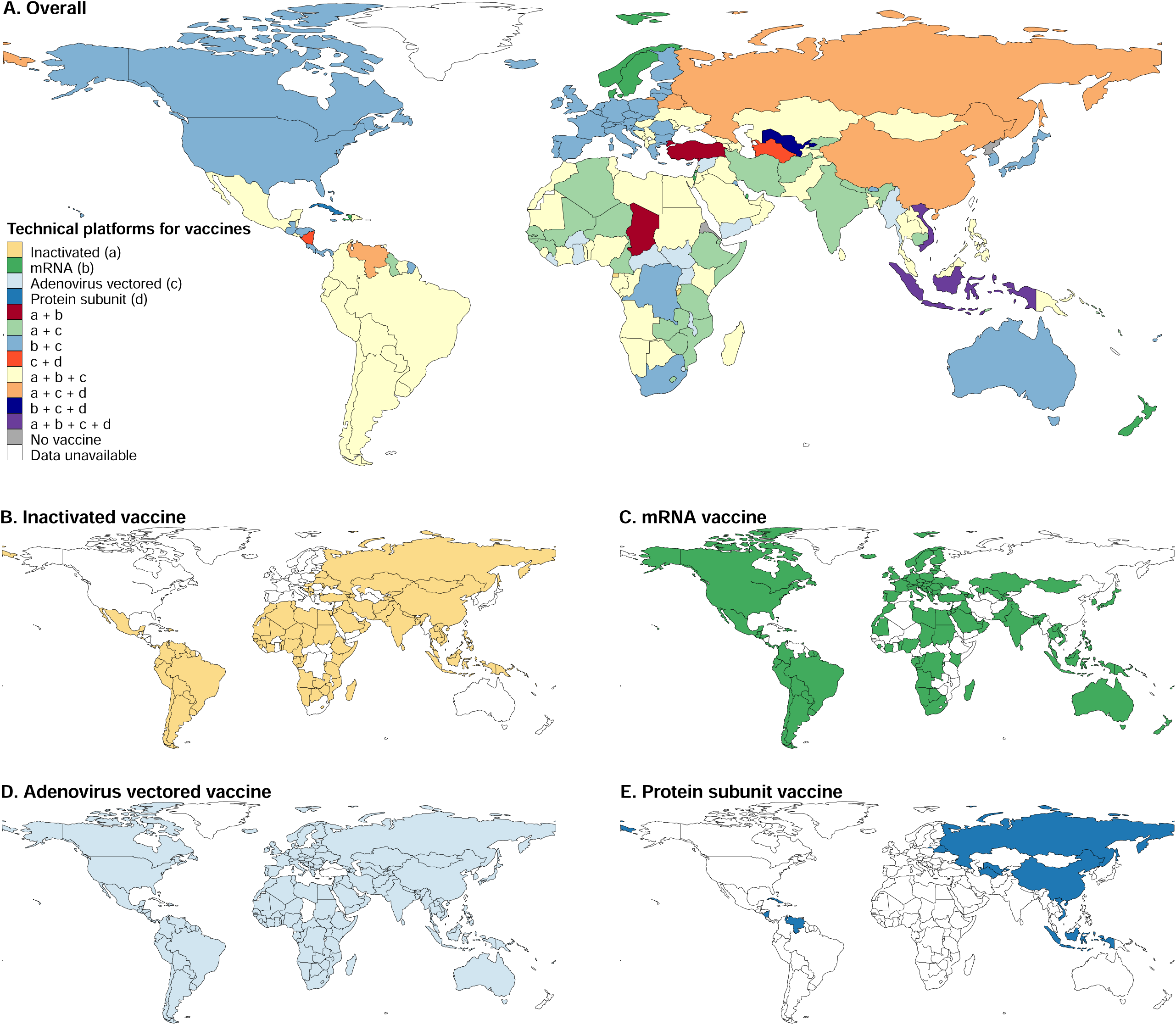
Technical platforms for vaccines being administered across countries. Geographic distribution of (A) overall technical platforms for vaccines, (B) inactivated vaccine, (C) mRNA vaccine, (D) adenovirus vectored vaccine, and (E) protein subunit vaccine administered across the globe. Since the conjugate vaccine was only used in three countries (Cuba, Iran and Nicaragua), it is not shown in the map.

Twelve different age groups were authorized worldwide for primary vaccination. Among 176 countries, vaccines were authorized for those 12 years and over in 94 (53.4%) countries and 18 years and over in 58 (33.0%) countries; five countries (Cambodia, Chile, Cuba, El Salvador and the United Arab Emirates) have approved vaccination for 2-, 3-, or 6-year-olds and above; Myanmar and Democratic Republic of the Congo has approved vaccination of people 55 years and older (Fig 2A). Additional doses are recommended in 41 countries, mainly targeting people at risk of serious illness (38 of 41 countries) (Fig 2B). Booster doses have been approved for use in 67 countries, but with wide variation in target population; 25 countries recommend booster doses for the general public (Fig 2C). Among 156 countries for which we could determine payment source, all provide COVID-19 vaccines to public free of charge, with exception of Singapore (not free for BBIBP-CorV vaccine) (Table S2).

**Figure 2.**
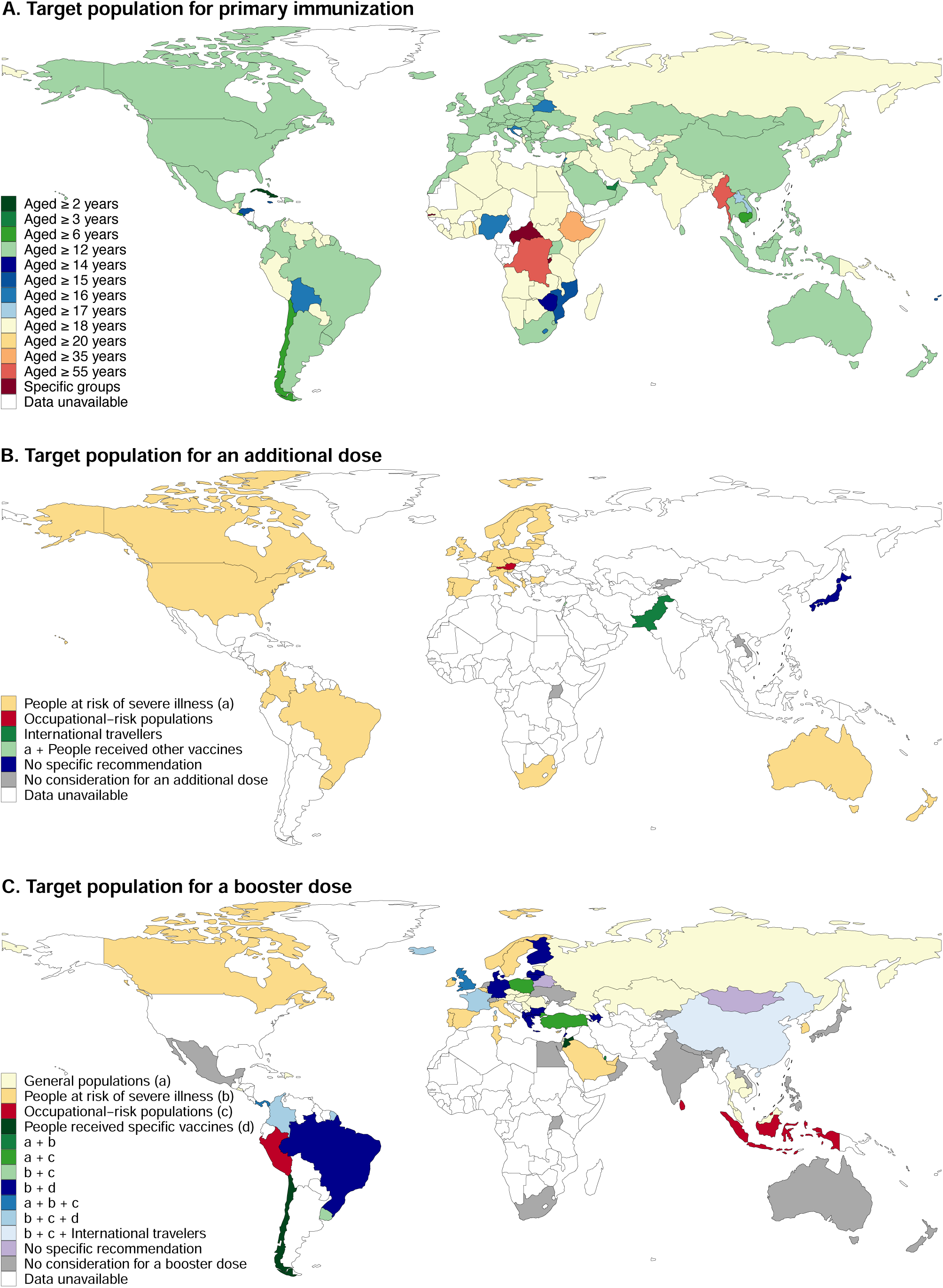
Target populations for primary immunization, and additional and booster doses. (A) Global distribution of target populations (mainly regulator-approved age groups) for primary vaccination; three countries vaccinated only people at risk (Central African Republic, Burundi, and Gambia). (B-C) Target populations for an additional dose and for a booster dose. People at risk of severe illness included elderly, residents in health centres, people with immunodeficiency disorders, people with autoimmune diseases receiving immunosuppressive therapy, people on dialysis or after organ transplantation, patients with onco-hematological diseases receiving treatment, and people with other comorbidities. Occupational-risk populations included workers at long-term care facilities, front-line health workers, and other front-line workers. People receiving specific vaccine types included people who receiving the one of the following vaccines: Janssen COVID-19 Vaccine, WBIP-CorV, and CoronaVac.

### Vaccine coverage

Vaccination data for 184 countries were available as of 7 October 2021; 116.5 doses have been administered per 100 target populations; 66.4% and 49.5% of target populations received at least one dose and full-schedule doses, respectively; 25.6% (46/180) and 10.4% (19/182) of countries have vaccinated more than two thirds of their population with at least one dose or full-schedule doses, respectively (Fig 3). Time needed to achieve one dose per 100 people varied between countries (Fig S3).

**Figure 3.**
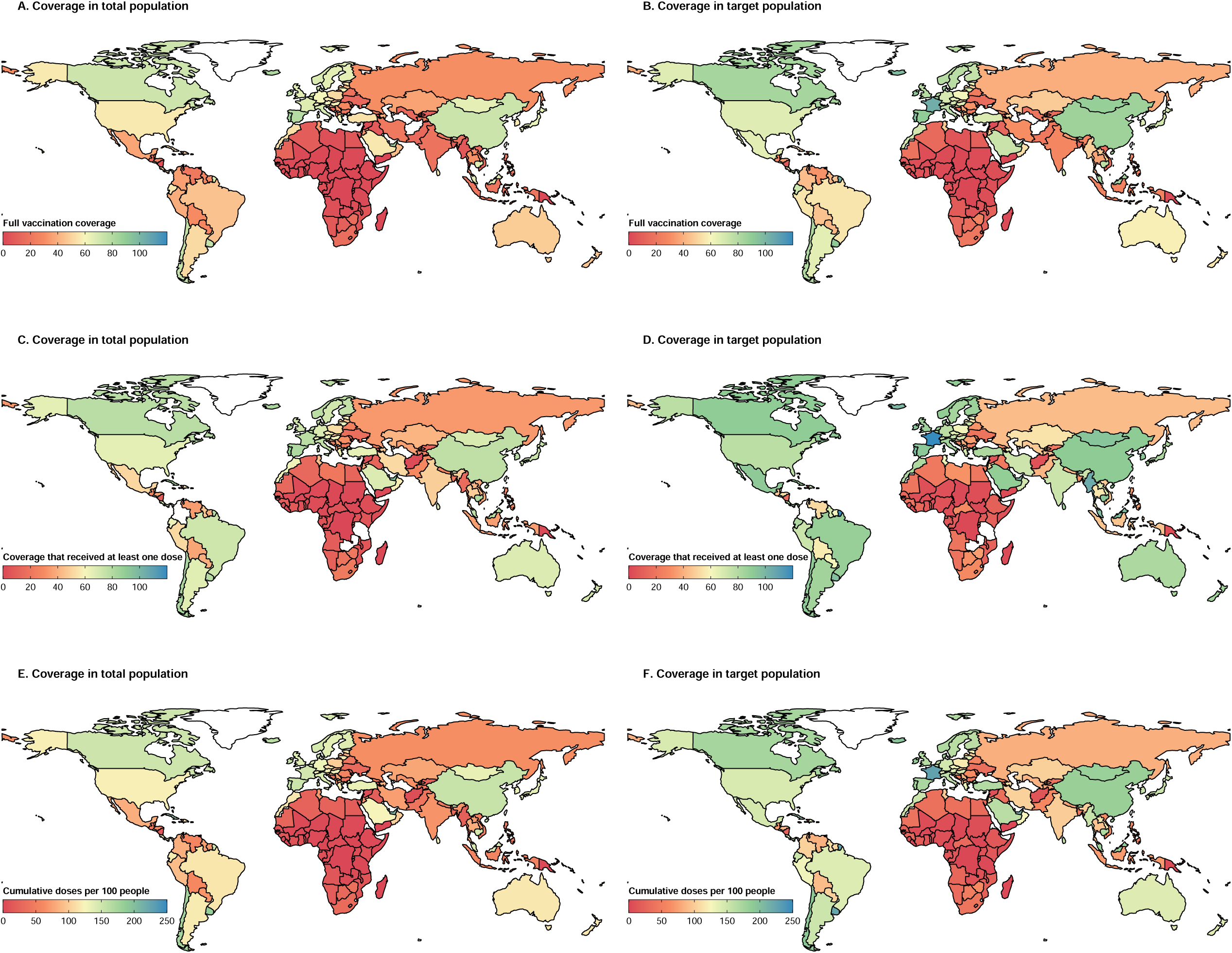
Vaccine coverage among total or target populations. Country-level full vaccine coverage among total populations (A) or target populations (B). Country-level proportion of people that received at least one dose among total populations (C) or target populations (D). Country-level cumulative doses per 100 people among total populations (E) or target populations (F). The white areas represent countries with no vaccination rollout or for which data are unavailable. The data shown here are as of October 7, 2021. Percentages of target populations that received at least one dose that exceed 100% may represent off-label use.

Highest full-dose target population coverage (77.2%) and doses administered (165.8 doses per 100 target populations) were reported in the WHO Western Pacific Region, followed by Europe (61.2%, 131.3 doses per 100 target populations), the Americas (60.3%, 138.3 doses per 100 target populations), South-East Asia (27.8%, 92.4 doses per 100 target populations), Eastern Mediterranean (27.1%, 68.5 doses per 100 target populations), and Africa (6.5%, 16.4 doses per 100 target populations) (Fig 3B, F). There was significant inter-country heterogeneity in full-dose coverage among target populations, ranging from 0% (Burundi) to more than 95.0% (San Marino, Malta, France, Iceland, Portugal) (Fig 3B), and cumulative doses per 100 target populations, ranging from 0 (Burundi) to 239.6 (Israel) (Fig 3F). In high-income countries, 82.8% of target populations have received at least one dose; in low-income countries, 21.2% of target populations have received at least one dose (Fig S4A). Countries selling or donating vaccine had higher coverage than those receiving vaccine (Fig S4C).

Among 22 countries reporting vaccine administration data by age group, the percent of people that received full-schedules was highest among seniors (people over 60 or 65 years old) (range: 33.6-100.6%), followed by middle-aged adults (18-60 or 18-65 years old) (32.6-94.6%), adolescents (12-17 years old) (1.0-95.8%), and children (0-11 years old) (0.01-22.8%) (Fig 4A). A similar pattern of vaccine distribution was also observed in one-or-more-dose coverage (Fig 4B). An exception was that the proportion of adolescents that received at least one dose in China was higher than for seniors receiving one or more doses.

**Figure 4.**
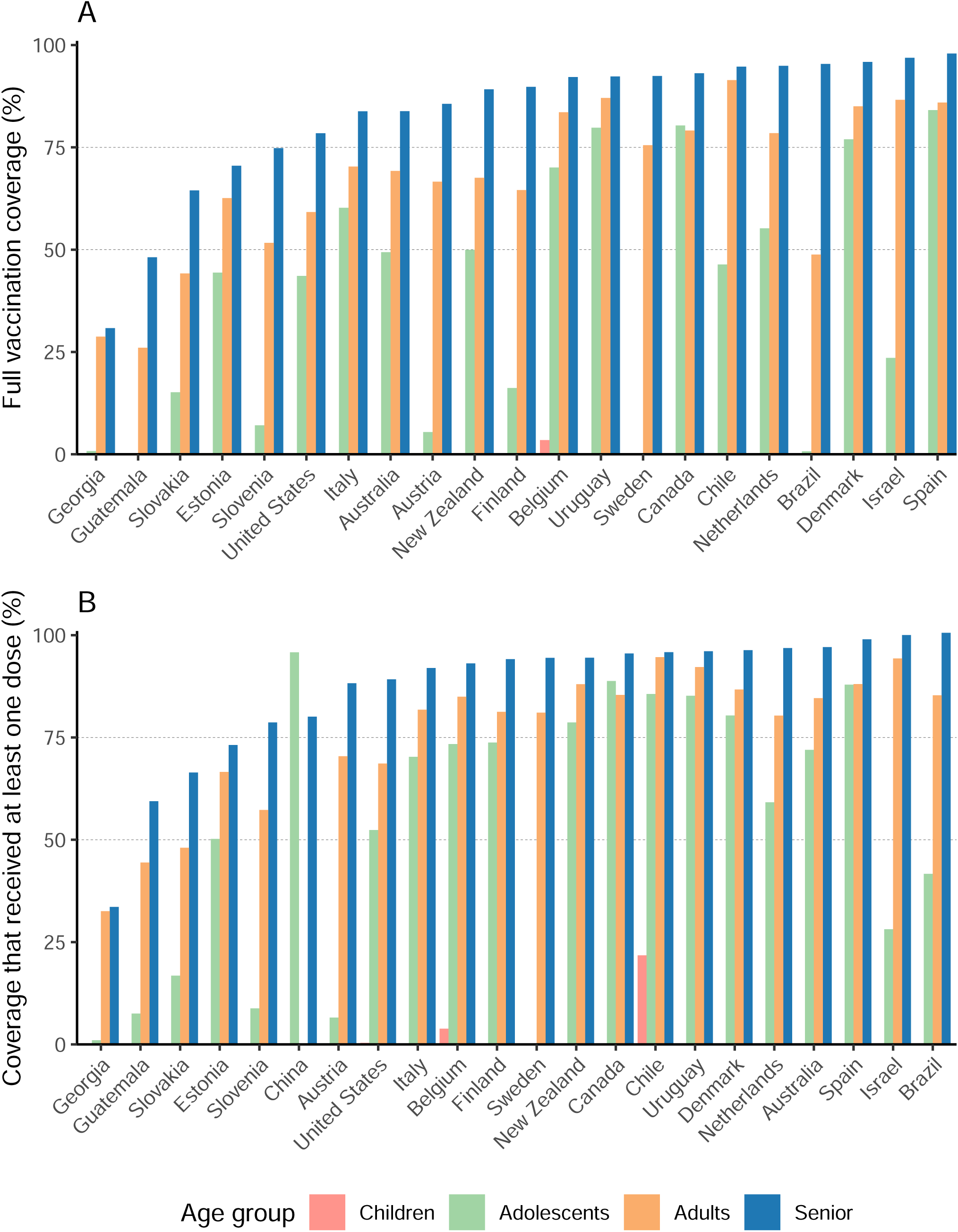
Vaccine coverage by age group. Proportion of age-specific populations that received full doses (A) and at least one dose (B) among total populations. The dividing line between age groups was not consistent across countries. Children refers to people aged 0-11 years old; adolescent refers to people aged 12-17 years old; adult refers to people aged 18-64 (Austria, Belgium, Chile, United States), 18-59 years old (Brazil, Canada, Estonia, Georgia, Guatemala, Iceland, Slovakia, Slovenia, Sweden, Netherland), 20-59 (Australia, Finland, Israel, Italy, Spain), or 20-64 (Denmark, New Zealand, Uruguay); and senior refers to people aged over 60 or 65 years old. Some countries only reported data for 0-17 or 0-19 year olds (Austria, Australia, Finland, Georgia, Israel, Slovenia, Slovakia, New Zealand, Uruguay), which is designated as the adolescent group. Adult data in China were unavailable.

### Influencing factors of vaccine coverage

Vaccine coverage was moderately associated with SDI, HAQ, and GDP per capita-PPP. Correlations between doses administered per 100 persons in the total population and the HAQ index (R^2^ = 0.58), SDI index (R^2^ = 0.56), and GDP per capita-PPP (R^2^ = 0.65) are shown in Figures 5A, C, E; correlations per 100 persons in the target population and HAQ index (R^2^ = 0.50), SDI index (R^2^ = 0.49), and GDP per capita-PPP (R^2^ = 0.56) are shown in Figures 5B, D, F. In general, countries with higher socio-demographic or health resource-related levels had higher coverage. As GDP per capita-PPP increased to approximately 50,000 dollars, coverage plateaued (Figures 5E-F).

**Figure 5.**
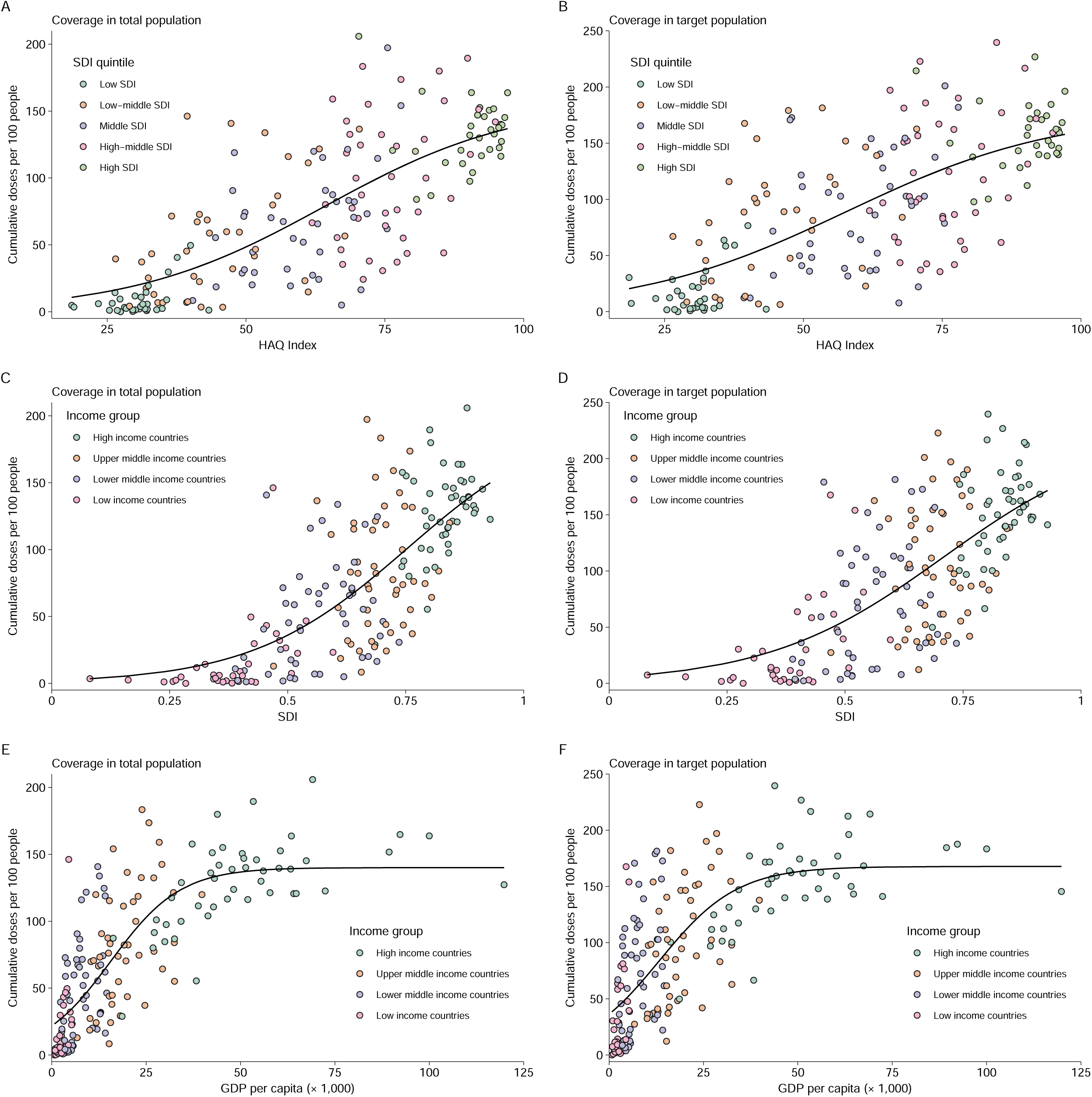
Correlations with vaccine coverage. Correlation of cumulative doses administered per 100 people among total populations or target populations with healthcare access and quality (HAQ) index (A-B), socio-demographic index (SDI) (C-D), GDP per capita (units: dollars) after adjusting for purchasing power parity (E-F). Solid line shows the nonlinear least squares’ fit.

### Demands of vaccine doses

The total estimated global demand was 5.54 billion doses to complete ongoing vaccination programs – 4.65 billion for primary immunization and 0.89 billion for additional/booster programs. The highest demand occurred in South-East Asia (1.51 billion doses), followed by Africa (0.98 billion doses), Europe (0.87 billion doses), Western Pacific (0.87 billion doses), the Americas (0.67 billion doses), and Eastern Mediterranean (0.64 billion doses) (Table S7). The total requirement for doses in low-income countries (0.78 billion doses) was similar to high-income countries (0.72 billion doses).

Global demand for primary immunization was 0.84 dose per individual in the target population, with regional-heterogeneity observed in the African (1.83), Eastern Mediterranean (1.31), South-East Asian (1.08), European (0.71), American (0.64), and Western Pacific (0.34) regions (Fig 6A). Estimated vaccine-dose demands at the country level are presented in the Appendix (Table S7). Additional and booster immunization policies increased demand to 0.96 per individual in the target population, as most additional/booster doses have not yet been administered to target populations (Fig 6B).

**Figure 6.**
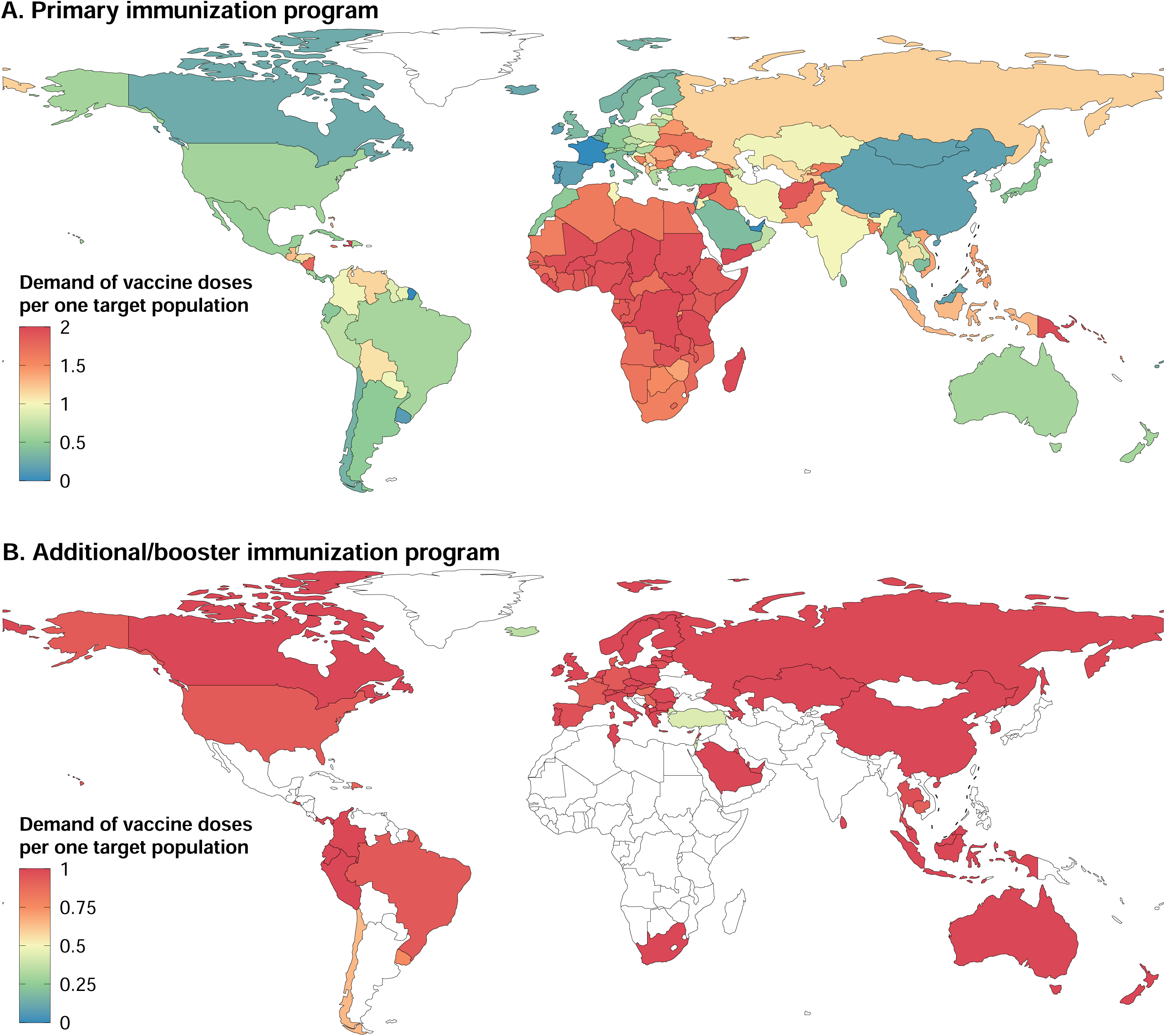
Current demand of vaccine dose per individual in the target population. (A) Demand of vaccine doses for primary immunization per individual in the target population. (B) Demand of vaccine dose for additional/booster immunization per individual in the target population. Target population is defined as those eligible for primary or additional/booster vaccination recommended by each country’s immunization policy.

## Discussion

Our study provides insight into the global landscape of COVID-19 vaccination policy, vaccine coverage, and demand of vaccines at this phase of vaccine rollout. Most countries have made COVID-19 vaccination a priority, and the 6.4 billion doses of COVID-19 vaccines administered around the world is a significant milestone in the response to the COVID-19 pandemic. With increases in vaccine production, the main target populations globally were adults aged 12 years and older, with some countries extending vaccination to children. However, we showed vast differences in vaccine coverage across countries, in which doses administered per capita in high-income countries was 8.8 times that of low-income countries. As a result, a huge imbalance in demand raises concerns of inequitable access to vaccines, even as production capacity increases.

Vaccination has shaped the epidemic curve of COVID-19, especially for severe outcomes^1^. However, we found a pattern similar to the dilemma of vaccine distribution during the 2009 H1N1 influenza pandemic in which high-income countries procured most vaccines, and access to vaccines remained inequitable^29,30^. Although vulnerable groups have not been effectively protected in some resource-poor countries, countries with high vaccine coverage have initiated booster programs based on the evidence that current COVID-19 vaccines provide sustained protection from severe outcomes and even from variants of concern, but antibody levels have waned^31^. This may further widen the gap of population immunity between countries. Disparities in coverage among countries may impede the global effort of building herd immunity to stop the pandemic. Slow vaccine rollouts in some low-income countries leave individuals vulnerable to emerging variants, and the expanding epidemic may further increase risks of new SARS-CoV-2 mutations^32^. Optimal allocation for limited supplies of vaccines to people at high risk or who have no immunity may save more lives and help contain the pandemic by inhibiting further evolution of variants. Although COVAX planned to procure and deliver at least 2 billion doses by the end of 2021^33^, only 367 million doses have been successfully allocated as of August 2021^21^.

Scaling up production capacity of current vaccines and developing more effective vaccines remain top priorities, especially since concerns over waning immunity and SARS-CoV-2 variants have led some countries to deploy extra vaccine doses. It may be feasible to use a booster dose with less antigen as a dose-sparing strategy that still provides adequate immune reponse^34,35^. With limited supplies of several vaccines, heterologous prime-boost regimens also should be considered because they appear to induce strong immune responses^36^. Research and development of one-dose, variant-specific, and broad-spectrum vaccines could curb the pandemic more effectively and efficiently. It is also vital to strengthen international coordination of development, manufacturing, and deployment, for example, by sharing knowledge and expertise, and providing useful guidance to build or improve production facility layouts and production lines^37^, enabling more countries to make vaccines.

In addition to challenges of production, affordability, and allocation, vaccine hesitancy is a key barrier for some countries with sufficient supplies to reach the expected vaccine coverage among target population^38^. Since vaccine hesitancy is not a singular problem, interventions should be implemented to build and sustain vaccine confidence through joint efforts by vaccine manufacturers, governments, and other parties to ensure safety and effectiveness of vaccines and provide timely disclosure of relevant information to the public^39^. Acceptance of COVID-19 vaccines may change over time as more robust evidence and monetary incentive policies emerge^40^.

Our univariate analysis showed that socioeconomic and health system-related factors might be predictors of vaccine coverage. Countries with a higher SDI and HAQ score were more likely to have more vaccine doses administered than those with lower scores. This finding may reflect that healthcare systems play a key role in vaccine rollout, because health-related resources and capabilities are necessary for vaccination campaigns. For example, inadequate equipment for temperature control and time lags between shipments and deployment of vaccines may impede vaccination progress in low-income countries. Arguably, widespread vaccine coverage needs to fit multiple efforts simultaneously, at global, national, and sub-national levels, including accessibility of vaccine, individual acceptance, and healthcare system requirements for vaccination^5,41^.

Planning COVID-19 vaccination programs is complex and arduous. Some concerns were raised during vaccination campaigns. Vaccination strategy for individuals previously infected with SARS-CoV-2 remains unclear, while the number of confirmed cases worldwide has reached 242 million, accounting for 3.1% of the total global population^1^. Interim evidence suggests that antibody levels in naturally-infected people persist for over one year^42,43^. Immune responses in individuals with mild previous infection who were given one dose of COVID-19 vaccine are higher than responses following full-schedule vaccination of people who had not been previously infected^44,45^. Therefore, a one-dose schedule for infected individuals might be acceptable to achieve the dual purpose of protecting populations and saving supplies. Certainly, more evidence is needed to guide future policies.

There are several limitations to this study. First, all data in our study were obtained from public sources. Therefore untimely, opaque, and language-restricted data disclosure limited data completeness and prediction of subpopulation immunity. Standardizing reporting of COVID-19 vaccination and increasing data sharing and transparency could promote progress of the global vaccination campaign. Second, estimates of population sizes with contraindications and immunosuppressing conditions were constrained because it is difficult to determine what proportions of these populations could be vaccinated. Some countries use vaccines off label, further complicating determination of the eligible population. Third, country vaccination policy will change with more real-world evidence and experience; vaccine demand in our study reflected only the current situation.

Our full picture of COVID-19 vaccination policy, coverage, and current demand in an ongoing epidemic deepens the understanding of this unprecedent vaccination effort. Disparity and inequity of vaccination rollout worldwide implies that susceptibility of unvaccinated populations in some countries may impede or reverse pandemic control, especially in the face of Delta and future variants. More countries and organizations should be involved in the global response to the pandemic, taking responsibility and providing leadership to overcome the complex challenges that lie ahead - financially, politically, and technically.

## Data Availability

All data used in this analysis will be available to others upon publication. Any additional information is available from the lead contact upon reasonable request.

## Contributors

H.Y. was responsible for its conception and design. W.Z., C.P., Q.W., Y.T., R.S., M.W., X.Z., Z.Z., G.Z., X.Y., N.L., F.H., S.Z., and T.Z. collected the data. Z.C., X.C., and C.P. did the analysis and prepared the figures. X.C., Y.T., R.S. and X.Z. prepared the tables. Z.C., W.Z., and Q.W. wrote the first draft. H.Y., A.S.A., and J.Y. made critical revision of the manuscript for important intellectual content. All authors contributed to review and revision and approved the final manuscript as submitted and agree to be accountable for all aspects of the work.

## Acknowledgments

We thank Dr. Lance Rodewald from Chinese Center for Disease Control and Prevention for his English language editing on this paper. This study was funded by Key Program of the National Natural Science Foundation of China (grant no. 82130093 to HJY) and the US National Institutes of Health (R01 AI135115 to ASA).

