## Supplementary Material for "Global diversity of policy, coverage, and demand of COVID-19 vaccines: a descriptive study"

#### COVID-19 vaccination policy

##### *Metrics included in the dataset of COVID-19 vaccination policy*

(1) **Authorization status**. Status includes licensure in use, emergency use authorization, conditional marketing, special access route, and recipients of COVAX or other countries.

(2) **Indications and contraindications**. People who could and should not get vaccinated against COVD-19, recommended by governments/health departments.

(3) **Additional/booster dose policy**. If the government/health department gives a clear definition of an additional/booster dose, we adopted it. Otherwise, we used the definition by the United States Centers for Disease Control and Prevention[1].

3.1 **Target population**. Individuals who are eligible for a COVID-19 vaccine additional/booster shot.

3.2 **Interval**. The time between the completion of primary immunization and additional/booster shots.

(5) **Whether provide at no cost to individual vaccine recipients.**

**Table S1. Authorization information of COVID-19 vaccines by technical platforms and country.**

| Vaccine | Authorization status | Use status | Country lists |
|---|---|---|---|
| **Adenovirus vectored vaccine** | | | |
| 1. Convidecia | Conditional marketing | In use | China |
| | Emergency Use Authorization | In use | Argentina, Chile, Ecuador, Indonesia, Malaysia, Mexico, Pakistan |
| 2. Covishield | Conditional marketing | Currently not in use | Canada |
| | Emergency Use Authorization | In use | Argentina, Bahrain, Bangladesh, Barbados, Bhutan, Botswana, Brazil, Cambodia, Cameroon, Comoros, Dominica, Egypt, Ghana, India, Iran, Laos, Madagascar, Maldives, Mauritius, Mexico, Morocco, Nepal, Nicaragua, Nigeria, Nigeria, Oman, Moldova, Saint Vincent and the Grenadines, Seychelles, South Africa, South Korea, Sri Lanka, Ukraine, Ukraine, Uzbekistan |
| | Recipients of COVAX or other countries | In use | Afghanistan, Angola, Ethiopia, Grenada, Guinea, Lesotho, Liberia, Malawi, Mali, Mauritania, Papua New Guinea, Saint Kitts and Nevis, Somalia, Suriname, Syria, Togo, Uganda, Yemen, Zambia, Trinidad and Tobago |
| 3. Janssen COVID-19 Vaccine | Conditional marketing / Emergency Use Authorization | Currently not in use | Denmark, Finland, Norway, Sweden, India |
| | Conditional marketing | In use | Austria, Bulgaria, Canada, Croatia, Cyprus, Czechia, Estonia, France, Germany, Greece, Iceland, Ireland, Italy, Latvia, Lithuania, Luxembourg, Malta, Netherlands, Romania, Slovakia, Slovenia, Spain |
| | Emergency Use Authorization | In use | Antigua and Barbuda, Bahamas, Bahrain, Bangladesh, Belgium, Belize, Bolivia, Botswana, Brazil, Cameroon, Chile, Colombia, Comoros, Cote d'Ivoire, Egypt, Ghana, Guinea, Hungary, Indonesia, Kenya, Kuwait, Laos, Lesotho, Libya, Madagascar, Malaysia, Marshall Islands, Mexico, Micronesia, Namibia, Nigeria, Oman, Philippines, Poland, Portugal, Rwanda, Saint Vincent and the Grenadines, Saudi Arabia, |

| | | | |
|---|---|---|---|
| | | | South Africa, South Korea, Sudan, Switzerland, Thailand, Togo, Tunisia, United Kingdom, United States, Vietnam, Zambia, Zimbabwe, Trinidad and Tobago |
| | Recipients of COVAX or other countries | In use | Afghanistan, Algeria, Benin, Central African Republic, Djibouti, Ethiopia, Gambia, Liberia, Malawi, Mali, Mauritania, Nepal, Palau, Papua New Guinea, Senegal, Sierra Leone, Somalia, South Sudan, Tajikistan, Tanzania, Yemen |
| 4. Sputnik Light | Emergency Use Authorization | In use | Belarus, Mongolia, Venezuela |
| | Licensure | In use | Russia |
| 5. Sputnik V | Emergency Use Authorization | Currently not in use | Maldives |
| | Emergency Use Authorization | In use | Albania, Algeria, Antigua and Barbuda, Argentina, Armenia, Azerbaijan, Bahrain, Bangladesh, Belarus, Bolivia, Bosnia and Herzegovina, Ecuador, Egypt, Gabon, Ghana, Guatemala, Guinea, Guyana, Honduras, Hungary, India, Iran, Iraq, Jordan, Kazakhstan, Kyrgyzstan, Libya, Mexico, Mongolia, Montenegro, Myanmar, Namibia, Nepal, Nicaragua, Nigeria, North Macedonia, Oman, Pakistan, Paraguay, Philippines, Moldova, Saint Vincent and the Grenadines, San Marino, Serbia, Seychelles, Slovakia, Sri Lanka, Syria, Tunisia, Turkmenistan, United Arab Emirates, Uzbekistan, Venezuela, Vietnam, Zimbabwe |
| | Licensure | In use | Russia |
| | Recipients of COVAX or other countries | In use | Angola |
| 6. Vaxzevria | Conditional marketing | Currently not in use | Denmark, Norway, Sweden |
| | Conditional marketing | In use | Austria, Bulgaria, Canada, Croatia, Cyprus, Czechia, Estonia, Finland, France, Germany, Greece, Iceland, Ireland, Italy, Latvia, Lithuania, Luxembourg, Malta, Netherlands, Romania, Slovakia, Slovenia, Spain, |

| | | | |
|---|---|---|---|
| | | | Vietnam |
| | Emergency Use Authorization | In use | Albania, Andorra, Antigua and Barbuda, Argentina, Armenia, Australia, Azerbaijan, Bahamas, Belgium, Belize, Benin, Bolivia, Bosnia and Herzegovina, Botswana, Brazil, Brunei, Burkina Faso, Cabo Verde, Chile, Colombia, Costa Rica, Cote d'Ivoire, Dominica, Dominican Republic, Ecuador, Egypt, El Salvador, Georgia, Guatemala, Guyana, Honduras, Hungary, Indonesia, Iraq, Japan, Kenya, Kuwait, Kyrgyzstan, Lebanon, Libya, Malaysia, Mexico, Mongolia, Montenegro, Namibia, Nauru, Nicaragua, Niger, North Macedonia, Oman, Pakistan, Panama, Paraguay, Peru, Philippines, Poland, Portugal, Saint Lucia, Saint Vincent and the Grenadines, Samoa, Saudi Arabia, Serbia, Thailand, Tunisia, Ukraine, United Arab Emirates, United Kingdom, Uruguay |
| | Recipients of COVAX or other countries | In use | Afghanistan, Algeria, Central African Republic, Democratic Republic of the Congo, Eswatini, Ethiopia, Fiji, Gambia, Guinea-Bissau, Jamaica, Jordan, Kiribati, Malawi, Mauritania, Mozambique, Papua New Guinea, Republic of the Congo, Rwanda, Sao Tome and Principe, Senegal, Sierra Leone, Solomon Islands, Somalia, South Sudan, Sudan, Tajikistan, Timor-Leste, Togo, Tonga, Tuvalu, Vanuatu, Zambia |
| **Conjugate vaccine** | | | |
| 1. Soberana 02 | Emergency Use Authorization | In use | Cuba, Iran, Nicaragua |
| **DNA vaccine** | | | |
| 1. ZyCoV-D | Emergency Use Authorization | Currently not in use | India |
| **Inactivated vaccine** | | | |
| 1. BBIBP-CorV | Conditional marketing | In use | China |
| | Emergency Use | In use | Antigua and Barbuda, Argentina, Armenia, Bahrain, Bangladesh, Barbados, Belarus, Belize, Bolivia, |

| | | | |
|---|---|---|---|
| | Authorization | | Brunei, Cambodia, Cameroon, Comoros, Dominica, Dominican Republic, Egypt, Equatorial Guinea, Gabon, Gambia, Georgia, Guyana, Hungary, Indonesia, Iran, Iraq, Jordan, Kenya, Kyrgyzstan, Laos, Lesotho, Madagascar, Maldives, Mauritania, Mauritius, Mongolia, Montenegro, Morocco, Namibia, Nepal, Niger, Nigeria, North Macedonia, Oman, Pakistan, Paraguay, Peru, Philippines, Republic of the Congo, Saudi Arabia, Senegal, Serbia, Seychelles, Sri Lanka, Suriname, Thailand, United Arab Emirates, Venezuela, Vietnam, Zimbabwe, Trinidad and Tobago |
| | Recipients of COVAX or other countries | In use | Afghanistan, Angola, Burundi, Chad, Cote d'Ivoire, Ethiopia, Guinea-Bissau, Kazakhstan, Kiribati, Lebanon, Mozambique, Papua New Guinea, Rwanda, Sierra Leone, Solomon Islands, Somalia, Sudan, Tanzania, Vanuatu, Zambia |
| | Special access route | In use | Singapore |
| 2. CoronaVac | Conditional marketing | In use | China |
| | Emergency Use Authorization | Currently not in use | Panama |
| | Emergency Use Authorization | In use | Albania, Azerbaijan, Bangladesh, Benin, Bosnia and Herzegovina, Botswana, Cambodia, Chile, Colombia, Djibouti, Dominican Republic, Ecuador, Egypt, El Salvador, Equatorial Guinea, Georgia, Guinea, Indonesia, Indonesia, Kazakhstan, Malaysia, Mexico, Nepal, North Macedonia, Oman, Pakistan, Paraguay, Philippines, Singapore, Thailand, Tunisia, Turkey, Ukraine, Uruguay, Venezuela, Zimbabwe |
| | Licensure | In use | Brazil |
| | Recipients of COVAX or other countries | In use | Algeria, Armenia, Libya, Mali, Moldova, Tajikistan, Timor-Leste, Togo |
| 3. Covaxin | Emergency Use Authorization | In use | Botswana, Comoros, Guyana, India, Iran, Mauritius, Mexico, Nepal, Paraguay, Philippines, Zimbabwe |

| | | | |
|---|---|---|---|
| 4. Covidful | Emergency Use Authorization | In use | China |
| 5. COVIran Barekat | Emergency Use Authorization | In use | Iran |
| 6. CoviVac | Licensure | In use | Russia |
| 7. FAKHRAVAC | Emergency Use Authorization | In use | Iran |
| 8. KCONVAC | Emergency Use Authorization | In use | China |
| 9. QazVac | Emergency Use Authorization | In use | Kazakhstan, Kyrgyzstan |
| 10. WBIP-CorV | Conditional marketing | In use | China |
| **mRNA vaccine** | | | |
| 1. Comirnaty | Conditional marketing | In use | Austria, Bulgaria, Canada, Croatia, Cyprus, Czechia, Denmark, Estonia, Finland, France, Germany, Greece, Iceland, Ireland, Italy, Latvia, Lithuania, Luxembourg, Malta, Monaco, Netherlands, Romania, San Marino, Slovakia, Slovenia, South Korea, Spain, Sweden |
| | Emergency Use Authorization | In use | Albania, Andorra, Argentina, Armenia, Australia, Azerbaijan, Bahamas, Bahrain, Bangladesh, Barbados, Belgium, Belize, Bhutan, Bolivia, Bosnia and Herzegovina, Botswana, Brazil, Brunei, Cabo Verde, Chile, Colombia, Cook Islands, Costa Rica, Democratic Republic of the Congo, Dominican Republic, Ecuador, Egypt, El Salvador, Gabon, Georgia, Guatemala, Honduras, Hungary, Indonesia, Iraq, Israel, Jamaica, Japan, Jordan, Kazakhstan, Kenya, Kuwait, Laos, Lebanon, Libya, Madagascar, Malaysia, Maldives, Marshall Islands, Mauritania, Mexico, Micronesia, Mongolia, Montenegro, Morocco, Namibia, New |

| | | | |
|---|---|---|---|
| | | | Zealand, Nigeria, Niue, North Macedonia, Norway, Oman, Pakistan, Panama, Papua New Guinea, Paraguay, Peru, Philippines, Poland, Portugal, Qatar, Moldova, Saint Kitts and Nevis, Saint Lucia, Saint Vincent and the Grenadines, Serbia, Singapore, South Africa, Sri Lanka, Suriname, Thailand, Tunisia, Turkey, Ukraine, United Arab Emirates, United Kingdom, United States, Uruguay, Vietnam, Trinidad and Tobago |
| | Licensure | In use | Saudi Arabia, Switzerland |
| | Recipients of COVAX or other countries | In use | Angola, Antigua and Barbuda, Benin, Chad, Cote d'Ivoire, Dominica, Grenada, Mauritius, Rwanda, Samoa, Sierra Leone, Sudan, Togo, Uzbekistan |
| 2. Spikevax | Conditional marketing | In use | Austria, Bulgaria, Canada, Croatia, Cyprus, Czechia, Denmark, Estonia, Finland, France, Germany, Greece, Iceland, Ireland, Italy, Latvia, Lithuania, Luxembourg, Malta, Netherlands, Romania, San Marino, Slovakia, Slovenia, Spain, Sweden |
| | Emergency Use Authorization | Currently not in use | India |
| | Emergency Use Authorization | In use | Argentina, Australia, Belgium, Botswana, Colombia, Democratic Republic of the Congo, Guatemala, Haiti, Honduras, Hungary, Indonesia, Israel, Japan, Kenya, Kuwait, Libya, Marshall Islands, Micronesia, Mongolia, Nigeria, Norway, Oman, Pakistan, Paraguay, Philippines, Poland, Portugal, Qatar, South Korea, Saint Vincent and the Grenadines, Saudi Arabia, Singapore, Sri Lanka, Suriname, Switzerland, Thailand, Ukraine, United Arab Emirates, United Kingdom, United States, Uzbekistan, Vietnam |
| | Recipients of COVAX or other countries | In use | Fiji, Palau, Republic of the Congo, Rwanda, Tajikistan, Tunisia |
| **Protein subunit vaccine** | | | |
| 1. Abdala | Emergency Use | In use | Cuba, Nicaragua, Venezuela, Vietnam |

| | | | |
|---|---|---|---|
| | Authorization | | |
| 2. EpiVacCorona | Emergency Use Authorization | In use | Turkmenistan |
| | Licensure | In use | Russia |
| | Recipients of COVAX or other countries | In use | Belarus |
| 3. Zifivax | Emergency Use Authorization | In use | China, Indonesia, Uzbekistan |

**Table S2. Self-payment Fee , target population, and contraindications recommended by regulatory agencies**

| Country | Payment | Target population | Contraindicated population | Data source |
|---|---|---|---|---|
| **Africa** | | | | |
| Algeria | Free | 18+ | No data | http://www.xinhuanet.com/english/2020-12/21/c_139605514.htm http://www.news.cn/english/2021-09/05/c_1310168528.htm |
| Angola | Free | 18+ | No data | https://www.macaubusiness.com/angola-free-covid-19-vaccinations-should-begin-in-feb-with-priority-for-over-40s-health-minister/ https://minsa.gov.ao/ao/ https://ao.usembassy.gov/covid-19-information/ |
| Benin | Free | 18+ | No data | https://documents1.worldbank.org/curated/en/936011623022343506/pdf/Project-Information-Document-Benin-COVID-19-second-Vaccines-Additional-Financing-to-COVID-19-Preparedness-and-Response-Project-P176562.pdf |
| Botswana | No data | 18+ | No data | https://fenyacovid.gov.bw/armready/ |
| Burkina Faso | Free | No data | No data | https://bf.usembassy.gov/u-s-citizen-services/covid-19-information/ |
| Burundi | No data | HCW; 60+; people with underlying conditions; people travelling abroad; any volunteers | No data | https://www.africanews.com/2021/10/19/burundi-begins-covid-19-vaccination-despite-hesitancy-among-its-people// |
| Cabo Verde | Free | 18+ | No data | https://reliefweb.int/report/cabo-verde/new-world-bank-support-equitable-access-covid-19-vaccines-cabo-verde |

| | | | | |
|---|---|---|---|---|
| | | | | https://cv.usembassy.gov/covid-19-information/ |
| Cameroon | No data | No data | No data | |
| Central African Republic | No data | HCW; elderly people and / or people with chronic diseases | No data | https://peacekeeping.un.org/en/national-covid-19-vaccination-campaign-officially-launched-central-african-republic |
| Chad | No data | No data | No data | |
| Comoros | No data | 18+ | No data | https://vaksiny.gov.mg/#/signin?from=/ |
| Cote d'Ivoire | Free | 18+ | No data | https://www.sante.gouv.ci/welcome/actualites/1192 |
| Democratic Republic of the Congo | No data | HCW; 55+; People suffering from serious health conditions such as kidney disease, high blood pressure or diabetes mellitus | No data | https://healthpolicy-watch.news/dr-congo-finally-launches-covid-19-vaccinations-after-concerns-about-astrazeneca/ |
| Equatorial Guinea | Free | No data | No data | https://gq.usembassy.gov/covid-19-information/ |
| Eritrea | No data | No data | No data | |
| Eswatini | No data | 12+ | No data | https://twitter.com/eswatinigovern1/status/1445737600732459017?lang=ar-x-fm |
| Ethiopia | No data | 35+; 18+ with health problems | People with a history of anaphylaxis to any component of the vaccine or an anaphylactic reaction following the first dose of this vaccine | https://www.moh.gov.et/ejcc/am/press-release-on-covid19 https://www.ephi.gov.et/images/novel_coronavirus/EPHI_PHEOC_COVID-19_Weekly_Bulletin_45_English_03132021.pdf |

| | | | | |
|---|---|---|---|---|
| Gabon | No data | No data | No data | |
| Gambia | Free | HCW; People with underlying medical condition; 65+ | No data | https://www.facebook.com/permalink.php?story_fbid=526763178764376&id=100866698020695 https://www.unicef.org/wca/press-releases/first-tranche-covid-19-vaccines-arrives-gambia-covax-initiative |
| Ghana | Free | 18+ | No data | http://fdaghana.gov.gh/news-media.php?page=78 http://www.xinhuanet.com/english/2021-08/17/c_1310130940.htm |
| Guinea | No data | No data | No data | |
| Guinea-Bissau | No data | No data | No data | |
| Kenya | Free | 18+ | No data | https://www.gavi.org/vaccineswork/ins-and-outs-kenyas-covid-19-vaccine-rollout-plan |
| Lesotho | No data | 16+ | No data | https://www.gov.ls/his-majesty-launches-covid-19-vaccine-roll-out/ |
| Liberia | Free | 18+ | No data | https://frontpageafricaonline.com/health/crusaders-for-peace-unicef-partner-to-end-covid-19-through-rapid-immunization/ https://lr.usembassy.gov/wp-content/uploads/sites/53/COVID-19-Vaccine-Fact-Sheet_FINAL_3Aug21.pdf |
| Madagascar | Free | 18+ | No data | https://mg.usembassy.gov/u-s-citizen-services/security-and-travel-information/covid-19-information/ |
| Malawi | Free | 18+ | No data | https://mg.usembassy.gov/u-s-citizen-services/security-and-travel |

| | | | | |
|---|---|---|---|---|
| | | | | -information/covid-19-information/ |
| Mali | No data | 18+ | No data | http://www.sante.gov.ml/index.php/actualites/communiques/item/6341-lutte-contre-la-covid-19-au-mali-835-200-doses-de-sinovac-pour-poursuivre-la-campagne-de-vaccination |
| Mauritania | Free | No data | No data | https://www.aa.com.tr/en/africa/mauritania-begins-covid-19-vaccination-campaign/2189322 |
| Mauritius | Free | 12+ | No data | https://health.govmu.org/Documents/Main%20Page/Corona/775_New%20Registration%20and%20Consent%20Form%20Covid-19%20Vaccination.pdf |
| Mozambique | No data | 15+ | Pregnant women | https://covid19.ins.gov.mz/vacina-covid-19/ https://covid19.ins.gov.mz/primeiro-ministro-lancou-esta-sexta-feira-o-plano-nacional-de-vacinacao/ |
| Namibia | Free | 18+ | No data | https://www.facebook.com/permalink.php?story_fbid=813016316038040&id=339157366757273 https://www.namibian.com.na/208360/archive-read/Namibias-Covid-19-vaccine-delayed |
| Niger | No data | 18+ | No data | https://ne.usembassy.gov/u-s-citizen-services/covid-19-information/ |
| Nigeria | Free | 16+ | People with allergic reactions to vaccine component | https://nphcda.gov.ng/faqs/ |
| Republic of the Congo | Free | No data | No data | https://docs.google.com/forms/d/e/1FAIpQLSc79P2xWViJpzwYs7rJ_n661seDoHEu-ZlH69qp7jdDkLq9Aw/viewform |
| Rwanda | Free | 18+ | No data | https://www.premiumtimesng.com/news/more-news/430797-rwa |

| | | | | |
|---|---|---|---|---|
| | | | | nda-offers-free-covid-19-vaccinations-to-citizens.html https://rbc.gov.rw/index.php?id=100&tx_news_pi1%5Bnews%5D=621&tx_news_pi1%5Bday%5D=22&tx_news_pi1%5Bmonth%5D=8&tx_news_pi1%5Byear%5D=2021&cHash=de0254720f9498593147d16c69f4cf8d |
| Sao Tome and Principe | No data | No data | No data | |
| Senegal | Free | 18+ | No data | https://sn.usembassy.gov/covid-19-information/ |
| Seychelles | Free | 12+ | No data | https://allafrica.com/stories/202108270089.html http://www.health.gov.sc/index.php/covid-19-vaccination-faqs/ |
| Sierra Leone | Free | 18+ | No data | https://dhse.gov.sl/wp-content/uploads/2021/09/PUBLIC-NOTICE-covid-113092021-1-1.pdf |
| South Africa | Free | 12+ | People with a history of severe allergic reaction to any ingredient in the vaccine; people who is allergic to polyethene glycol (PEG) should not get the Pfizer vaccine, as it is one of the components; people who had a severe allergic reaction after the first dose should not get the second dose of that vaccine | https://www.sahpra.org.za/wp-content/uploads/2021/04/SAHPRA-Registration-with-conditions-for-the-Covid-19-vaccine-Janssen-Media-release-statement_final.pdf https://www.iol.co.za/sunday-tribune/news/children-over-12-now-eligible-to-receive-covid-vaccine-58327f81-c0a3-49de-b4b0-097c605fa3aa https://www.gov.za/coronavirus/faqs/vaccine |
| South Sudan | Free | 18+ | No data | https://www.afro.who.int/news/south-sudan-receives-first-batch-covid-19-vaccines-through-covax-facility https://www.afro.who.int/news/south-sudan-receives-its-first-cons |

| | | | | |
|---|---|---|---|---|
| | | | | ignment-johnson-johnson-covid-19-vaccines-through-covax |
| Tanzania | Free | 18+ | No data | https://www.afro.who.int/news/united-republic-tanzania-receives-first-covax-shipment<br>https://tz.usembassy.gov/covid-19-information/ |
| Togo | Free | 20+ | No data | https://vaccin.covid19.gouv.tg/faq/<br>https://vaccin.covid19.gouv.tg/strategie-vaccinale/ |
| Uganda | No data | 12+ | No data | http://www.xinhuanet.com/english/africa/2021-07/26/c_1310086931.htm |
| Zambia | Free | 18+ | No data | https://www.usaid.gov/zambia/press-releases/sep-3-2021-united-states-provides-additional-67-million-covid-assistance<br>https://www.afro.who.int/news/zambia-launches-covid-19-vaccination |
| Zimbabwe | Free | 14+ | No data | https://www.aa.com.tr/en/africa/zimbabwe-free-vaccination-to-begin-thursday/2147237<br>https://apnews.com/article/lifestyle-africa-health-zimbabwe-coronavirus-pandemic-3e0b007ccccf08ae285710250daa84be |
| **Americas** | | | | |
| Antigua and Barbuda | Free | 12+ | Pregnant / breastfeeding women; persons who have severe allergic reactions to ingredients of the vaccine | https://www.facebook.com/investingforwellness/posts/1262969224143049<br>https://vaccineantiguabarbuda.com/faq/ |
| Argentina | Free | 12+ | People with an allergic reaction to vaccine components or first dose; an acute SARS-CoV-2 | https://www.argentina.gob.ar/coronavirus/vacuna/preguntas-frecuentes#7 |

| | | | | |
|---|---|---|---|---|
| | | | infection; pre-vaccination pregnancy | https://www.argentina.gob.ar/coronavirus/vacuna/preguntas-frecuentes#22 |
| Bahamas | Free | 12+ | No data | https://www.bahamas.gov.bs/wps/portal/public<br>https://vax.gov.bs/ |
| Barbados | Free | 12+ | No data | https://gisbarbados.gov.bb/blog/pfizer-vaccine-now-available-to-the-general-public/ |
| Belize | No data | 12+ | No data | https://www.facebook.com/Belizehealth/posts/2767419013556496 |
| Bolivia | Free | 16+ | No data | https://www.unidoscontraelcovid.gob.bo/index.php/vacunas/<br>https://www.unidoscontraelcovid.gob.bo/index.php/2021/10/07/ministerio-de-salud-coordinara-con-educacion-para-inmunizar-contra-el-covid-19-a-menores-de-edad-entre-16-y-17-anos/ |
| Brazil | Free | 12+ | People with allergic action to the first dose or any vaccine component | https://translate.googleusercontent.com/translate_f#21 |
| Canada | Free | 12+ | No data | https://www.canada.ca/en/public-health/services/diseases/coronavirus-disease-covid-19/vaccines/how-vaccinated.html |
| Chile | Free | 6+ | People with a known history of severe acute allergy (anaphylaxis) | https://www.gob.cl/yomevacuno/<br>https://www.gob.cl/yomevacuno/preguntasfrecuentes/ |
| Colombia | Free | 12+ | No data | https://www.como.gov/covidvaccine/#elementor-toc__heading-anchor-5<br>https://www.rcnradio.com/colombia/mayores-de-70-anos-podran-recibir-el-refuerzo-anticovid-de-su-vacuna-inicial-de-pfizer-o |

| | | | | |
|---|---|---|---|---|
| Costa Rica | Free | 12+ | No data | https://www.ccss.sa.cr/arc/covid19/Manual_procedimientos_vacunacion_COVID.pdf |
| Cuba | Free | 2+ | No data | https://salud.msp.gob.cu/actualizacion-de-la-vacunacion-en-el-marco-de-los-estudios-de-los-candidatos-vacunales-cubanos-y-la-intervencion-sanitaria/ https://salud.msp.gob.cu/actualizacion-de-la-vacunacion-en-el-marco-de-los-estudios-de-los-candidatos-vacunales-cubanos-y-la-intervencion-sanitaria/ |
| Dominica | No data | 12+ | No data | http://news.gov.dm/news/5337-pfizer-vaccines-arrive-in-dominica |
| Dominican Republic | No data | 12+ | No data | https://covid-19pharmacovigilance.paho.org/pfizer-biontech |
| Ecuador | No data | 12+ | No data | https://ec.usembassy.gov/covid-19-information-ecu-2/ https://www.coronavirusecuador.com/2021/09/inicio-vacunacion-a-poblacion-de-12-a-15-anos-en-coordinacion-entre-los-ministerios-de-salud-y-educacion/ |
| EI Salvador | Free | 6+ | No data | https://www.presidencia.gob.sv/la-poblacion-meta-a-vacunar-contra-covid-19-incremento-a-5-9-millones-con-la-inclusion-de-los-nuevos-grupos-etarios/ |
| Grenada | Free | 12+ | People with a severe allergic reaction to any ingredient in a COVID-19 vaccine | https://covid19.gov.gd/faq-vaccination/ https://caribbean.loopnews.com/content/grenada-begins-its-pfizer-vaccine-roll-out-soon |

| | | | | |
|---|---|---|---|---|
| Guatemala | No data | 18+ | People with a fever; people with serious illness; pregnant women | https://www.mspas.gob.gt/component/jdownloads/category/890-plan-nacional-de-vacunaci%C3%B3n-contra-la-covid-19.html?Itemid=-1 |
| Guyana | Free | 18+ | No data | https://www.health.gov.gy/ |
| Haiti | Free | No data | No data | https://ht.usembassy.gov/covid-19-information/ |
| Honduras | Free | 15+; 12-15 with disability | People with allergic reaction to the first dose or vaccine component; people with a fever | http://www.salud.gob.hn/site/index.php/component/edocman/resumen-ejecutivo-ampliacion-vii-campana-de-vacunacion-covid-19-v0709 http://www.salud.gob.hn/site/index.php/component/edocman/resumen-ejecutivo-ampliacion-vii-campana-de-vacunacion-covid-19-v0709 |
| Jamaica | Free | 15+; 12-15 with underlying conditions | No data | https://vaccination.moh.gov.jm/frequently-asked-questions/ |
| Mexico | Free | 12+ | People with allergic reaction to vaccine component; people received blood transfusion or monoclonal antibodies | http://vacunacovid.gob.mx/wordpress/preguntas-frecuentes/ |
| Nicaragua | No data | No data | No data | |
| Panama | Free | 12+ | No data | http://www.minsa.gob.pa/noticia/traslado-de-insumos-y-personal-clave-en-la-estrategia-de-vacunacion-en-el-12-1 |
| Paraguay | Free | 18+ | No data | https://www.gov.uk/foreign-travel-advice/paraguay/coronavirus https://www.vacunate.gov.py/index-plan-vacunacion.html |
| Peru | Free | 18+; 12-18 with type III | No data | https://doh.gov.ph/vaccines/know-your-vaccines |

| | | | | |
|---|---|---|---|---|
| | | obesity, diabetes mellitus, cancer, Down syndrome, rare and orphan diseases | | |
| Saint Kitts and Nevis | Free | 12+ | People who are allergic to substances in the vaccine; pregnant and breastfeeding women | https://covid19.gov.kn/covid-19-vaccination/ https://www.sknis.gov.kn/2021/08/13/pfizer-covid-19-vaccines-will-offer-protection-to-the-federations-children-12-years-and-up-minister-of-health-byron-nisbett/ |
| Saint Lucia | Free | 12+ | No data | https://suntci.com/people-in-st-lucia-can-now-register-online-for-covid-vaccine-p5886-135.htm http://socialtransformation.govt.lc/news/covid-19-vaccination-update2 |
| Saint Vincent and the Grenadines | Free | No data | No data | https://www.gov.uk/foreign-travel-advice/st-vincent-and-the-grenadines/coronavirus |
| Suriname | Free | 18+ | People with proven severe allergies to any of the components of the vaccine; pregnant women | https://laatjevaccineren.sr/vragen/#vaccineren |
| United States | Free | 12+ | People with allergic reactions to vaccine component | https://www.fda.gov/media/144413/download https://www.cdc.gov/coronavirus/2019-ncov/vaccines/different-vaccines/Pfizer-BioNTech.html |
| Uruguay | Free | 12+ | People with allergic reactions to the first dose or vaccine component | https://www.gub.uy/ministerio-salud-publica/comunicacion/publicaciones/preguntas-frecuentes-vacunacion-covid-19/sobre-vacuna |

| | | | | |
|---|---|---|---|---|
| | | | | s/casos-indicados |
| Venezuela | Free | 18+ | No data | https://reporting.unhcr.org/sites/default/files/Panama%20operational%20update%20April%202021.pdf |
| Trinidad and Tobago | Free | 12+ | People with allergic reactions to vaccine component | https://health.gov.tt/covid-19/covid-19-vaccine/faqs |
| **Eastern Mediterranean** | | | | |
| Afghanistan | No data | 18+ | No data | https://www.adb.org/sites/default/files/linked-documents/55012-001-sd-03.pdf |
| Bahrain | Free | 12+; 3-11 with respiratory diseases, heart diseases, diabetes mellitus, obesity, cancer, Down syndrome and birth defects | People should not have allergies to any of the vaccine's components; women should not be pregnant or planning on getting pregnant, or lactating women | https://healthalert.gov.bh/uploads/dhep3ew1_l2l.pdf https://healthalert.gov.bh/uploads/sf3iuypu_oby.pdf https://www.cvdvaccine-bh.com/files/V6_6-2-Final-121120-EUA_Full-Prescribing-Info_HCP-Fact-Sheet-Pfizer-BioNTech-COVID-19-vaccine-14-1-2021.pdf https://www.nhra.bh/Media/Announcement/MediaHandler/GenericHandler/documents/Announcements/NHRA_News_MoH%20Circular_Inactivated%20COVID19%20Vaccine%20(Snopharm)_20201215n.pdf |
| Djibouti | No data | 18+ | No data | https://www.aa.com.tr/en/africa/djibouti-receives-more-covid-19-jabs-as-vaccination-drive-underway/2307470 |
| Egypt | Free | 18+ | No data | https://www.egypttoday.com/Article/1/103826/Egyptians-non-Egyptians-receive-COVID-vaccine-for-free-Health-Ministry https://www.thenationalnews.com/mena/egypt-opens-vaccine-regi |

| | | | | |
|---|---|---|---|---|
| | | | | stration-for-all-citizens-over-18-1.1186403 |
| Iran | No data | No data | No data | |
| Iraq | No data | 18+ | No data | https://www2.hse.ie/screening-and-vaccinations/covid-19-vaccine/get-the-vaccine/deciding-on-vaccination-for-12-to-15-year-olds/ |
| Jordan | Free | 12+ | People with allergic reactions to vaccine component, first dose, another vaccine; pregnant or breastfeeding women | https://corona.moh.gov.jo/ar/page/1061/CoronaVaccineQuestions |
| Kuwait | Free | 12+ | No data | http://www.xinhuanet.com/english/2021-07/19/c_1310069039.htm |
| Lebanon | Free | 16+ | People with an anaphylactic shock after receiving a previous dose of this vaccine, or a severe allergic reaction to any component of this vaccine | https://www.familyfirsthealth.org/lebanon-covid-19-vaccine-clinic<br>https://www.moph.gov.lb/userfiles/files/Prevention/COVID-19%20Vaccine/COVID-19%20Vaccine%20FAQ%20-%20EN%20-22-3-%202021.pdf<br>https://www.moph.gov.lb/userfiles/files/Prevention/COVID-19%20Vaccine/COVID-19%20Vaccine%20FAQ%20-%20EN%20-22-3-%202021.pdf |
| Libya | No data | 18+ | Pregnant and breastfeeding women | https://www.eservices.ly/ |
| Morocco | Free | 12+ | People with allergic reactions to vaccine component or the first dose; people allergic to other vaccine; pregnant or breastfeeding women; people with an infectious disease in the acute phase | https://www.africanews.com/2021/09/01/covid-19-morocco-begins-inoculating-children-over-12-years-old//<br>https://liqahcorona.ma/fr/page-je-minforme-sur-le-vaccin#209<br>https://liqahcorona.ma/fr/questions#faq |

| | | | | |
|---|---|---|---|---|
| Oman | Free | 12+ | No data | https://www.thearabianstories.com/2021/08/15/covid-19-free-vaccination-begins-for-oman-expats-in-al-dhahirah-governorate-1/ https://om.usembassy.gov/u-s-citizen-services/covid-19-information/ |
| Pakistan | Free | 12+ | No data | https://ncoc.gov.pk/covid-vaccination-en.php |
| Qatar | Free | 12+ | People with allergic reactions to vaccine component; pregnant women | https://covid19.moph.gov.qa/EN/Covid19-Vaccine/Pages/FAQ.aspx |
| Saudi Arabia | Free | 12+ | People seriously infected with SARS-CoV-2 | https://www.moh.gov.sa/en/eServices/Pages/Covid19-egistration.aspx https://articlesen.covid19awareness.sa/COVID-19-Vaccine-FAQs |
| Somalia | Free | 18+ | No data | https://reliefweb.int/report/somalia/race-against-time-boost-covid-19-vaccine-uptake-somalia |
| Sudan | Free | 18+ | People with allergic reaction to the first dose; people with a fever | https://www.facebook.com/FMOH.SUDAN/posts/2986575494948742 https://www.unicef.org/sudan/stories/covid-19-vaccination-sudan |
| Syria | Free | 18+ | No data | http://www.xinhuanet.com/english/2021-05/06/c_139926698.htm http://www.emro.who.int/syria/news/update-on-covid-19-vaccination-in-syria-22-september-2021.html |
| Tunisia | Free | 18+ | No data | https://reliefweb.int/report/tunisia/tunisia-receives-first-batch-covid-19-vaccines-through-covax-facility https://reliefweb.int/report/tunisia/tunisia-receives-first-batch-covid-19-vaccines-through-covax-facility |

| | | | | |
|---|---|---|---|---|
| United Arab Emirates | Free | 3+ | People with allergic reactions to vaccine component; pregnant women | https://u.ae/en/information-and-services/justice-safety-and-the-law/handling-the-covid-19-outbreak/vaccines-against-covid-19-in-the-uae. https://www.wam.ae/en/details/1395302961623 |
| Yemen | Free | No data | No data | https://www.thenationalnews.com/gulf-news/yemenis-urged-to-take-covid-19-vaccine-as-first-astrazeneca-shipment-arrives-1.1194914 |
| **Europe** | | | | |
| Albania | Free | 18+ | No data | http://shendetesia.gov.al/fushata-e-vaksinimit-shqiperia-buzeqesh/ |
| Andorra | Free | 12+ | No data | https://www.salut.ad/preinscripcio-vacuna-covid19/ |
| Armenia | Free | 18+ | If you have a history of severe allergic reactions to any ingredients of a COVID-19 vaccine; if you are currently sick or experiencing symptoms of COVID-19. | https://www.unicef.org/armenia/en/stories/everything-you-need-know-about-covid-19-vaccination-armenia#Question1 |
| Austria | Free | 12+ | Seriously allergic to first dose | https://www.sozialministerium.at/Corona-Schutzimpfung/Corona-Schutzimpfung---Fachinformationen.html |
| Azerbaijan | Free | 12+ | People with uncontrolled epilepsy; patients receiving immunosuppressive therapy; pregnant women | http://sehiyye.gov.az/xeberler/3418-azrbaycan-respublikasnn-shiyy-nazirliyi-cbari-tibbi-sorta-zr-dvlt-agentliyi-v-tibbi-razi-blmlrini-daretm-birliyinin-mlumati.html http://health.gov.az/xeberler/3477-azrbaycan-respublikas-shiyy-nazirliyinin-mlumati.html |
| Belarus | Free | 16+ | Immunosuppressive therapy | https://cms.law/en/int/expert-guides/cms-expert-guide-to-vaccine- |

| | | | | |
|---|---|---|---|---|
| | | | | compensation-regimes/belarus http://minzdrav.gov.by/ru/dlya-spetsialistov/rekomendatsii-po-vaktsinatsii-protiv-covid-19.php |
| Belgium | Free | 12+ | No data | https://www.info-coronavirus.be/en/vaccination/#faq |
| Bosnia and Herzegovina | No data | No data | No data | |
| Bulgaria | Free | 12+ | People who have allergic reactions to vaccine component | https://btvnovinite.bg/bulgaria/vaksina-sreshtu-covid-19-za-deca-interesat-ot-roditelite-zasega-e-plah.html?fbclid=IwAR20JTPCzPxeHkEHl7NARofgzgmHPmiuucuUrjbAGh7iWdCAnI7yl8CKxow https://coronavirus.bg/bg/vaccinations/faq |
| Croatia | Free | 16+ | People with an immediate allergic reaction to any other vaccine or injection therapy | https://www.koronavirus.hr/o-covidu/892 https://www.koronavirus.hr |
| Cyprus | No data | 12+ | No data | https://www.pio.gov.cy/coronavirus/uploads/06092021_vaccination3doseEN.pdf |
| Czechia | Free | 12+ | People with allergic reaction to vaccine component | https://covid.gov.cz/en/situations/information-about-vaccine/vaccine-indication-selected-cohorts |
| Denmark | Free | 12+ | People with a severe allergic reaction to the first shot or any ingredient of the vaccine; people with a fever | https://www.sst.dk/da/corona/Vaccination/Saadan-bliver-du-vaccineret https://www.sst.dk/-/media/Udgivelser/2021/Corona/Vaccination/Notater/Vidensbank-til-almen-praksis-og-vaccinationscentre.ashx?la=da&hash=A5DF7E41671F7370E23EE58D9124B5935FA55 |

| | | | | |
|---|---|---|---|---|
| | | | | D3D |
| Estonia | Free | 12+ | People with a severe allergic reaction to the first shot or any ingredient of the vaccine; people with a fever; people suffering from severe frailty syndrome, are in a very bad general condition or nearing the end of their life | https://www.kriis.ee/en/vaccination-plan-and-risk-groups https://kkk.kriis.ee/en/faq/covid-19-vaccination/vaccination-plan-and-risk-groups |
| Finland | Free | 12+ | People with allergic reactions to the first dose or vaccine component | https://thl.fi/fi/web/infektiotaudit-ja-rokotukset/rokotteet-a-o/koronavirusrokotteet-eli-covid-19-rokotteet-ohjeita-ammattilaisille https://thl.fi/en/web/infectious-diseases-and-vaccinations/what-s-new/coronavirus-covid-19-latest-updates/vaccines-and-coronavirus/suitability-of-covid-19-vaccines-for-various-groups |
| France | Free | 12+ | People with allergic reactions to the first dose or vaccine component; people with thrombocytopenia and coagulation disorders, Capillary leak syndrome; 12-17 with developed pediatric multisystem inflammatory syndrome | https://vaccination-info-service.fr/Les-maladies-et-leurs-vaccins/COVID-19 |
| Georgia | Free | 18+ | Pregnant women | https://www.provax.ge/Content/files/en_Vaccination_plan.pdf https://www.provax.ge/en/ |
| Germany | Free | 12+ | People with allergies to components of the COVID-19 vaccines; people with a fever | https://www.bundesgesundheitsministerium.de/coronavirus/faq-covid-19-impfung.html#c21988 https://www.rki.de/SharedDocs/FAQ/COVID-Impfen/gesamt.html |

| | | | | |
|---|---|---|---|---|
| Greece | Free | 12+ | People with a severe allergic reaction to the first shot or any ingredient of the vaccine | People with a severe allergic reaction to the first shot or any ingredient of the vaccine |
| Hungary | Free | 12+ | No data | https://koronavirus.gov.hu/gyik https://www.nnk.gov.hu/index.php/koronavirus-tajekoztato/936-gyakori-kerdesek-valaszok-a-vakcinarol-pfizer-biontech-vakcina http://www.xinhuanet.com/english/europe/2021-06/11/c_1310001374.htm |
| Iceland | Free | 12+ | People with a history of severe allergies | https://www.lyfjastofnun.is/frettir/bolusetning-barna-og-unglinga-12-15-ara/ https://www.lyfjastofnun.is/covid-19/comirnaty-biontech-pfizer-2/ https://www.landlaeknir.is/um-embaettid/greinar/grein/item44019/vaccination-against-covid-19 |
| Ireland | Free | 12+ | No data | https://www2.hse.ie/screening-and-vaccinations/covid-19-vaccine/get-the-vaccine/deciding-on-vaccination-for-12-to-15-year-olds/ |
| Israel | Free | 12+ | People with serious allergic reactions; people with a fever | https://govextra.gov.il/ministry-of-health/covid19-vaccine/en-covid19-vaccination-information/ |
| Italy | Free | 12+ | People with allergic reactions to the first dose | https://www.aifa.gov.it/en/domande-e-risposte-su-vaccini-covid-19 |
| Kazakhstan | Free | 12+ | No data | https://egov.kz/cms/en/articles/health_care/Vakcinaciya-protiv-koronavirusnoy-infekcii- https://primeminister.kz/en/news/press/deti-ot-12-let-i-starshe-sm |

| | | | | |
|---|---|---|---|---|
| | | | | ogut-poluchit-vakcinu-pfizer-tolko-na-dobrovolnoy-osnove-i-s-soglasiya-roditeley-a-coy-2782049 |
| Kyrgyzstan | Free | 18+ | People with allergic reaction to vaccine component; people with a fever; people with an acute attack of chronic disease; people with epilepsy; pregnant or breastfeeding women | http://med.kg/ru/novosti/5345-kak-zashchitit-lyudej-s-osobymi-potrebnostyami-ot-sovid-19-delat-li-vaktsinatsiyu.html http://med.kg/ru/vaktsinatsiya/5037-vaktsinatsiya-lits-s-nevrologicheskimi-zabolevaniyami.html |
| Latvia | Free | 12+ | People with a severe allergic reaction in the past after receiving other vaccines or medications | https://www.spkc.gov.lv/lv/manavakcinalv https://www.spkc.gov.lv/lv/visparigi-jautajumi-par-vakcinam |
| Lithuania | Free | 12+ | People with a fever | https://koronastop.lrv.lt/lt/duk/vakcinacija-nuo-covid-19/vakcinacija-nuo-12-metu-amziaus |
| Luxembourg | Free | 12+ | No data | https://covid19.public.lu/fr/vaccination/infovaxx.html |
| Malta | Free | 12+ | People with a history of serious allergy (anaphylaxis) following a vaccine or injectable medication | https://deputyprimeminister.gov.mt/en/health-promotion/covid-19/Pages/frequently-asked-questions.aspx https://deputyprimeminister.gov.mt/en/health-promotion/covid-19/Pages/travel.aspx |
| Moldova | Free | 18+ | No data | http://vaccinare.gov.md/permitted-vaccines |
| Monaco | Free | 12+ | No data | https://covid19.mc/en/fight-against-coronavirus/vaccination/qui-peut-se-faire-vacciner-contre-la-covid19/ https://covid19.mc/lutter-contre-la-covid-19/vaccination/ |
| Montenegro | Free | 12+ | No data | https://www.covidodgovor.me/me/cesta-pitanja |
| Netherlands | Free | 12+ | No data | https://www.government.nl/topics/coronavirus-covid-19/dutch-vaccination-programme |

| | | | | |
|---|---|---|---|---|
| North Macedonia | Free | 12+ | No data | http://zdravstvo.gov.mk/sq/jane-aplikuar-1-435-392-doza-te-vaksines-ndersa-jane-rivaksinuar-667-739-qytetare/ |
| Norway | Free | 12+ | People with allergic reactions to the first dose; people with a fever | https://www.fhi.no/publ/brev/informasjonsbrev-09.09.2021---gjennomforing-av-vaksinasjon-med-3.-dose/ |
| Poland | Free | 12+ | People with allergic reactions to the first dose or vaccine component; people with immunodeficiency; pregnant or breastfeeding women | https://www.gov.pl/web/szczepimysie/narodowy-program-szczepien-przeciw-covid-19<br>https://www.gov.pl/web/szczepimysie/jak-sie--zaszczepic<br>https://www.gov.pl/web/szczepimysie/pytania-i-odpowiedzi |
| Portugal | Free | 12+ | No data | https://covid19.min-saude.pt/perguntas-frequentes/<br>https://covid19.min-saude.pt/pedido-de-agendamento/<br>https://covid19.min-saude.pt/meta-alcancada-85-da-populacao-tem-a-vacinacao-completa/ |
| Romania | Free | 12+ | No data | https://vaccinare-covid.gov.ro/vaccinuri-autorizate/<br>https://apnews.com/article/europe-romania-coronavirus-pandemic-health-ecd443fc19c966e9b07f7b14cf88c830 |
| Russia | Free | 18+ | People with allergic reaction to vaccine component; people with a fever; people with an acute attack of chronic diseases; pregnant or breastfeeding women | https://static-0.minzdrav.gov.ru/system/attachments/attaches/000/054/706/original/%D0%9E%D1%82%D0%B2%D0%B5%D1%82%D1%8B_%D0%BF%D0%BE_%D0%B2%D0%B0%D0%BA%D1%86%D0%B8%D0%BD%D0%B0%D1%86%D0%B8%D0%B8_COVID_19-19.02.2021-Red.pdf |
| San Marino | Free | 12+ | People with allergic reaction;<br>People taking anticoagulant therapy | https://vaccinocovid.iss.sm/faq<br>https://vaccinocovid.iss.sm/vaccinazione-anticovi-fascia%2012-1 |

| | | | | |
|---|---|---|---|---|
| | | | | 5%20anni |
| Serbia | Free | 12+ | People with fever | https://www.france24.com/en/video/20210326-go-to-serbia-for-a-free-covid-vaccine-country-offers-jabs-to-foreigners https://vakcinacija.gov.rs/vakcine-protiv-covid-19-u-srbiji/ https://borgenproject.org/covid-19-vaccinations-in-serbia/ https://vakcinacija.gov.rs/vakcine-protiv-covid-19-u-srbiji/ |
| Slovakia | Free | 12+ | No data | https://www.slovenskoproticovidu.sk/sk/vsetko-o-ockovani/vakciny/viac-o-vakcine-comirnaty-od-vyrobcov-pfizer-biontech |
| Slovenia | Free | 12+ | People with severe allergy to the ingredients of the vaccine; people with a fever; people with thrombosis syndrome with thrombocytopenia after the first vaccination with Vaxzevria | https://www.nijz.si/sites/www.nijz.si/files/uploaded/priporocila_za_cepljenje_proti_covid_uskl_psc_apr_2021.pdf https://www.cepimose.si/cepljenje-proti-covidu-19/pogosta-vprasanja-in-odgovori/ |
| Spain | Free | 12+ | People with a history of having had severe allergic reactions (e.g., anaphylaxis) to any component of the vaccine | https://www.vacunacovid.gob.es/preguntas-y-respuestas/deben-vacunarse-los-ninos-y-las-ninas-y-la-poblacion-adolescente https://www.vacunacovid.gob.es/preguntas-y-respuestas |
| Sweden | Free | 12+ | No data | https://www.folkhalsomyndigheten.se/smittskydd-beredskap/utbrott/aktuella-utbrott/covid-19/vaccination-mot-covid-19/fragor-och-svar-om-vaccination-mot-covid-19/ https://www.1177.se/en/other-languages/other-languages/covid-19/vaccin-engelska/#section-133829 |
| Switzerland | Free | 12+ | No data | https://www.swissmedicinfo.ch/ShowText.aspx?textType=FI&lang=DE&authNr=68225 |

| | | | | |
|---|---|---|---|---|
| Tajikistan | Free | 18+ | No data | https://apa.az/en/xeber/cis-countries-news/tajikistan-declares-mandatory-covid-19-vaccination-353147 https://eurasianet.org/dashboard-vaccinating-eurasia-june |
| Turkey | Free | 12+ | People with allergic reactions to the first dose or vaccine component | https://covid19asi.saglik.gov.tr/EN-78316/frequently-asked-questions.html https://www.hurriyetdailynews.com/erdogan-urges-parents-teachers-to-get-covid-19-vaccine-167672 https://www.dailysabah.com/turkey/turkey-lowers-covid-19-vaccination-age-to-15-starts-fourth-doses/news |
| Turkmenistan | Free | 18+ | No data | https://tdh.gov.tm/en/post/25815/objectives-of-increasing-national-health-system%E2%80%99s-potential-discussed https://www.barrons.com/news/uzbekistan-certifies-russia-s-sputnik-vaccine-for-mass-use-01613559912 https://www.garda.com/crisis24/news-alerts/508931/turkmenistan-authorities-tighten-covid-19-countermeasures-in-ashgabat-as-of-aug-2-update-16 https://tdh.gov.tm/en/post/25911/turkmenistan-registers-vaccines-prevention-infectious-diseases |
| Ukraine | Free | 12+ | People with any contraindications or allergies to any vaccine; people with an acute illness and body temperature above 38.5ºC | https://vaccination.covid19.gov.ua/faq https://www.kyivpost.com/ukraine-politics/ukraine-allows-children-over-12-to-be-vaccinated-with-pfizer.html |
| United | Free | 12+ | No data | https://www.gov.uk/government/publications/regulatory-approval |

| | | | | |
|---|---|---|---|---|
| Kingdom | | | | -of-pfizer-biontech-vaccine-for-covid-19/information-for-uk-recipients-on-pfizerbiontech-covid-19-vaccine |
| Uzbekistan | Free | No data | No data | |
| **South-East Asia** | | | | |
| Bangladesh | No data | 18+ | No data | https://www.facebook.com/opmbs/posts/from-the-statement-the-astrazeneca-vaccine-will-be-administered-to-eligible-baha/762781414443838/ |
| Bhutan | No data | 12+ | People with a severe allergic reaction to the first shot or any ingredient of the vaccine; pregnant or breastfeeding women | https://drive.google.com/file/d/1UMlrlI-HvD5x1q99NHi50LoMP-_qjaIA/view https://www.gov.bt/covid19/03-03-21-information-series-on-covid-19-vaccines-moh/ |
| India | Free | 18+ | People with a severe allergic reaction to the first shot or any ingredient of the vaccine; pregnant or breastfeeding women; people with severe illness | https://www.mohfw.gov.in/vaccinationbooklet/#page/1 https://www.mohfw.gov.in/covid_vaccination/vaccination/faqs.html#about-the-vaccine |
| Indonesia | Free | 12+ | No data | https://www.kemkes.go.id/article/view/21070200001/phase-3-vaccination-begins-targeting-vulnerable-communities-and-children-of-12-17-years-old.html |
| Maldives | Free | 12+ | People with a fever | http://health.gov.mv/Uploads/Downloads//Informations/Informations(362).pdf https://covid19.health.gov.mv/vaccination/?c=0 https://raajje.mv/104861 |
| Myanmar | Free | 55+; people with | No data | https://www.mohs.gov.mm/page/17483 |

| | | | | |
|---|---|---|---|---|
| | | disabilities; members of the armed groups and their families from ethnic armed groups; displaced communities and temporary communities; persons with chronic and incurable diseases | | |
| Nepal | Free | 18+ | No data | https://reliefweb.int/sites/reliefweb.int/files/resources/Focused%20COVID-19_Media%20Monitoring_September%2020%2C%202021.pdf |
| North Korea | No data | No data | No data | |
| Sri Lanka | Free | 12+ | No data | http://www.epid.gov.lk/web/index.php?option=com_content&view=article&id=131:topics-recent&Itemid=487&lang=en http://www.epid.gov.lk/web/index.php?option=com_content&view=article&id=131:topics-recent&Itemid=487&lang=en |
| Thailand | Free | 12+ | No data | https://thainews.prd.go.th/en/news/detail/TCATG210727104313781 https://www.bangkokpost.com/thailand/general/2180323/students-aged-12-18-to-get-pfizer-jabs-later-this-month |
| Timor-Leste | Free | 18+ | No data | https://reliefweb.int/report/timor-leste/timor-leste-launches-covid- |

| | | | | |
|---|---|---|---|---|
| | | | | 19-vaccination-campaign |
| **Western Pacific** | | | | |
| Australia | Free | 12+ | No data | https://www.health.gov.au/initiatives-and-programs/covid-19-vaccines/getting-vaccinated-for-covid-19 |
| Brunei | Free | 12+ | No data | http://www.moh.gov.bn/Shared%20Documents/COVID-19%20Vaccine/FAQs%20General%20Covid%2019%20Vaccine_04032021.pdf<br>https://www.thestar.com.my/aseanplus/aseanplus-news/2021/10/20/brunei-to-vaccinate-children-aged-12-17<br>http://www.moh.gov.bn/Shared%20Documents/COVID-19%20Vaccine/Brunei%20Darussalam%20Vaccination%20Strategy%20V2.pdf |
| Cambodia | Free | 6+ | No data | Cambodia to Administer Booster Shots to General Public from Mid-October – Cambodia News Service<br>http://www.xinhuanet.com/english/asiapacific/2021-04/09/c_139869140.htm |
| China | Free | 12+ | Those who are allergic to the ingredients of the vaccine or who have history of allergy to the same type of vaccine; history of serious allergy to vaccines (such as acute allergic reactions, angioedema, breathing difficulty); people with uncontrolled epilepsy and other serious neurological diseases; | http://www.nhc.gov.cn/wjw/hygq/202104/8e62004e41d648d5a084b3fb7bf098ea.shtml<br>http://english.nmpa.gov.cn/2021-04/01/c_608350.htm |

| | | | | |
|---|---|---|---|---|
| | | | patients with fever, or acute diseases, or during acute attacks of chronic diseases, or patients with uncontrolled severe chronic diseases; women during pregnancy | |
| Cook Islands | Free | 12+ | No data | https://covid19.gov.ck/sites/default/files/2021-09/Approval%20given%20for%2012-15%20year%20old%20to%20be%20offered%20vaccine%202.pdf |
| Fiji | Free | 15+ | People with allergic reactions to the first dose | https://www.health.gov.fj/covid-19-vaccination-campaign/ https://www.health.gov.fj/children-15-17/ https://www.health.gov.fj/covid-vaccine/vaccine-faqs/ |
| Japan | Free | 12+ | People with allergic reactions to vaccine component; people with a fever. | https://www.mhlw.go.jp/stf/covid-19/vaccine.html https://www.cov19-vaccine.mhlw.go.jp/qa/receive/ https://www.mhlw.go.jp/content/000759294.pdf |
| Kiribati | Free | 18+ | No data | https://www.mhms.gov.ki/single.php?id=26 |
| Laos | Free | 17+ | No data | http://www.xinhuanet.com/english/asiapacific/2021-08/10/c_1310118766.htm https://www.thestar.com.my/aseanplus/aseanplus-news/2021/09/18/laos-health-ministry-advises-covid-19-vaccine-jabs-for-pregnant-women-and-older-students |
| Malaysia | Free | 12+ | People with allergic reaction to vaccine component; people with immunodeficiency | https://www.npra.gov.my/easyarticles/images/users/1047/Frequently-Asked-Questions-FAQ-about-Comirnaty-Covid-19-Vaccine.pdf |

| | | | | |
|---|---|---|---|---|
| Marshall Islands | No data | 18+ | No data | https://www.rnz.co.nz/international/pacific-news/436910/marshalls-leads-pacific-s-covid-19-vax-charg |
| Micronesia | Free | 12+ | No data | https://hsa.gov.fm/fsm-covid-19-vaccination/ https://gov.fm/index.php/component/content/article/35-pio-articles/news-and-updates/430-covid-19-vaccines-now-available-for-all-eligible-citizens-18-in-all-fsm-states |
| Mongolia | Free | 12+ | No data | https://moh.gov.mn/news/ |
| Nauru | No data | 18+ | No data | http://naurugov.nr/media/146514/nauru_bulletin__03_9jul2021__228_.pdf |
| New Zealand | Free | 12+ | People with a history of anaphylaxis to any component or previous dose of mRNA-CV | https://www.health.govt.nz/our-work/immunisation-handbook-2020/5-coronavirus-disease-covid-19 https://www.health.govt.nz/our-work/immunisation-handbook-2020/5-coronavirus-disease-covid-19 |
| Niue | Free | 12+ | No data | https://covid19.govt.nz/iwi-and-communities/translations/niuean/ |
| Palau | No data | 12+ | No data | https://covid19.govt.nz/iwi-and-communities/translations/niuean/the-covid-19-vaccine/vaccine-basics/ |
| Papua New Guinea | No data | 18+ | No data | https://postcourier.com.pg/chinese-vaccine-to-be-gazetted/ |
| Philippines | Free | 12+ | People with allergic reactions to vaccine component or the first dose | https://doh.gov.ph/vaccines https://doh.gov.ph/press-release/DOH-VACCINATION-AMONG-CHILDREN-AGED-12-17-TO-START-WITH-COMORBIDITIES-AS-PART-OF-A3-GROUP |

| | | | | |
|---|---|---|---|---|
| Samoa | Free | 12+ | People with allergic reactions to vaccine component; people with a fever; people taking anticoagulant therapy; people with immunodeficiency; pregnant women; 85+ (optional) | https://vaccinocovid.iss.sm/faq https://www.samoaobserver.ws/category/samoa/90307 |
| Singapore | Free | 12+ | No data | https://www.gov.sg/features/covid-19-vaccination |
| Solomon Islands | Free | 18+ | No data | https://solomons.gov.sb/roll-out-of-2nd-dose-of-astrazeneca-vaccine-now-underway-in-honiara/ |
| South Korea | No data | 18+ | People with allergic reactions to the first dose or vaccine component; pregnant women; people with acute symptoms such as fever (37.5℃ or higher) | http://english.seoul.go.kr/covid/covid-19-vaccination-guideline/ |
| Tonga | Free | 18+ | No data | https://matangitonga.to/2021/08/24/tonga-fully-vaccinates-43-eligible-people-over-18?fbclid=IwAR3KFE6s_Brg8uj0JWH-YhmejAg1fdrlv-2nh95NOLu_uL510fdPSfOCeT4 |
| Tuvalu | No data | 18+ | No data | https://www.facebook.com/AusHCfnfu/posts/750425468961962 |
| Vanuatu | No data | 18+ | No data | https://covid19.gov.vu/index.php/vaccination/information |
| Vietnam | Free | 12+ | No data | https://en.vietnamplus.vn/ministry-grants-conditional-approval-of-pfizer-biontech-vaccine/202977.vnp http://news.chinhphu.vn/Home/Viet-Nam-plans-to-vaccinate-adolescents-aged-1218-against-COVID19-this-month/202110/45761.vgp |

Abbreviation: 18+, people aged 18 years and above; HCW, healthcare workers

**Table S3. Recommendation on the use of an additional or booster dose of COVID-19 vaccine**

| Country | Additional dose | Notes for additional dose | Booster dose | Notes for booster dose | Data source |
|---|---|---|---|---|---|
| **Africa** | | | | | |
| Andorra | Yes | **Indication**: People with immunodeficiency; people on dialysis; people receiving a transplant | Yes | **Indication**: Vulnerable people like the elderly; residents in health centers | https://www.govern.ad/comunicats/item/13197-s-inicia-l-administracio-de-la-tercera-dosi-voluntaria-als-residents-a-centres-sociosanitaris-de-gent-gran |
| Mauritius | No data | | Yes | **Indication:** people receiving Sinopharm<br>**Interval**: 120 d | https://govmu.org/EN/newsgov/SitePages/2021/Vaccination-campaign-for-a-third-booster-Sinopharm-dose-to-kickstart-on-23-September-2021.aspx |
| South Africa | Yes | **Indication**: people with immunosuppression | Under discussion | | https://www.sanews.gov.za/south-africa/covid-19-booster-shot-immunocompromised-south-africans |
| Uganda | No policy | | No policy | | |
| **Americas** | | | | | |
| Brazil | Yes | **Indication**: People with immunosuppression<br>**Interval**: 28 d | Yes | **Indication**: 60+; HCW<br>**Interval**: 180 d | https://www.gov.br/saude/pt-br/coronavirus/vacinas/NTDoseReforo.pdf<br>https://www.reuters.com/world/americas/brazil-provide-covid-19-booster-shots-all-people-over-60-years-old-2021-09-28/ |
| Canada | Yes | **Indication**: People with moderate to severe immunocompromision | Yes | **Indication**: Long-term care residents; seniors living in other congregate settings<br>**Interval**: 180 d | https://www.canada.ca/en/public-health/services/immunization/national-advisory-committee-on-immunization-naci/statement-september-10-2021-additional-dose-covid-19-vaccine-immunocompromised-following-1-2-dose-series.html<br>https://www.canada.ca/en/public-health/services/immunization |

| | | | | | |
|---|---|---|---|---|---|
| | | | | | /national-advisory-committee-on-immunization-naci/statement-september-28-2021-booster-dose-long-term-care-residents-seniors-living-other-congregate-settings.html?hq_e=el&hq_m=2190695&hq_l=1&hq_v=e48b482e2d |
| Chile | No data | | Yes | **Indication:** People receiving Sinovac | https://www.bloomberg.com/news/articles/2021-08-05/chile-rolls-out-covid-19-booster-campaign-that-mixes-vaccines |
| Colombia | Yes | **Indication**: People with immunosuppression | Yes | **Indication:** 65+; 18+ with underlying conditions; pregnant women; 18+ at high-risk exposure of SARS-CoV-2; 18+ living in nursing home | https://www.como.gov/covidvaccine/#elementor-toc__heading-anchor-5 |
| Dominican Republic | No data | | Yes | **Indication**: 12+ | https://twitter.com/RaquelPenaVice/status/1408186389204590596 |
| Ecuador | Yes | **Indication**: people with weak immune systems | No data | | https://www.reuters.com/world/americas/ecuador-give-immune-weakened-people-third-covid-19-vaccine-2021-08-17/ |
| EI Salvador | No data | | Yes | **Indication**: 18+<br>**Interval**: 120 d | https://www.presidencia.gob.sv/el-salvador-supero-el-de-aplicaciones-de-la-tercera-dosis-de-la-vacuna-anticovid-19/ |
| Mexico | No data | | No policy | | |
| Panama | Yes | **Indication**: people with immunosuppression | Yes | **Indication**: HCW; 55+; people in nursing home; security forces; 18-54 at high | https://www.reuters.com/world/americas/panama-give-immunocompromised-people-third-covid-19-vaccine-shot-2021-09-22/ |

| | | | | | |
|---|---|---|---|---|---|
| | | | | risk of exposure; 18-54 with underlying conditions<br>**Interval**: 180 d | http://www.minsa.gob.pa/noticia/comunicado-ndeg-596 |
| Peru | No data | | Yes | **Indication**: HCW | https://www.youtube.com/watch?v=NiqPRcPo_1A |
| United States | Yes | **Indication**: People with immunosuppression<br>**Interval**: 28 d | Yes | **Indication**: 65+; 18+ who live in long-term care settings; 18+ who have underlying medical conditions; 18+ who work in high-risk settings; 18+ who live in high-risk settings<br>**Interval**: 180 d | https://www.cdc.gov/coronavirus/2019-ncov/vaccines/booster-shot.html<br>https://www.fda.gov/news-events/press-announcements/fda-issues-emergency-use-authorization-third-covid-19-vaccine |
| Uruguay | Yes | **Indication**: People with immunosuppression | Yes | **Indication**: People receiving Sinovac; 18+<br>**Interval**: 90 d | https://www.gub.uy/ministerio-salud-publica/comunicacion/noticias/administracion-dosis-refuerzo-vacuna-contra-covid-19-inmunodeprimidos<br>https://www.gub.uy/uruguaysevacuna |
| **Eastern Mediterranean** | | | | | |
| Bahrain | No data | | Yes | **Indication:** 18+<br>**Interval**: 180 d | https://healthalert.gov.bh/en/category/vaccine |
| Egypt | No data | | Under discussion | | |
| Jordan | No data | | Yes | **Indication**: People receiving Sinopharm | https://petra.gov.jo/Include/InnerPage.jsp?ID=36412&lang=en&name=en_news |

| | | | | | |
|---|---|---|---|---|---|
| Kuwait | No data | | Yes | **Indication**: 18+ | https://cov19vaccine.moh.gov.kw/SPCMS/CVD_19_Vaccine_Booster_Registration.aspx |
| Lebanon | Yes | **Indication**: People with immunosuppression; people receiving Sinopharm | Yes | **Indication**: 75+; frontline HCW | https://www.moph.gov.lb/en/Pages/127/55387/recommendations-of-the-national-committee-for-covid-19-vaccine-on-the-administration-of-the-third-do |
| Oman | No data | | Under discussion | | |
| Pakistan | Yes | **Indication**: 12+ international travelers | No data | | https://ncoc.gov.pk/covid-vaccination-en.php |
| Qatar | No data | | Yes | **Indication:** 50+; people with chronic conditions which increase their risk for severe COVID-19 infection; people with immunosuppression | https://covid19.moph.gov.qa/EN/Covid19-Vaccine/Pages/FAQ.aspx<br>https://www.gub.uy/ministerio-salud-publica/comunicacion/noticias/se-abre-agenda-terceras-dosis-18-59-anos |
| Saudi Arabia | No data | | | **Indication:** 60+<br>**Interval**: 240 d | https://english.alarabiya.net/coronavirus/2021/09/27/Saudi-Arabia-will-provide-third-booster-COVID-19-vaccine-dose-for-over-60-years-old |
| Tunisia | No data | | Yes | **Indication**: 50+ with underlying conditions | |
| United Arab Emirates | No data | | Yes | **Indication:** 60+; people with underlying condition; 18+ who receiving long-term | https://www.wam.ae/en/details/1395302977389 |

| | | | | | |
|---|---|---|---|---|---|
| | | | | health care<br>**Interval**: 180 d | |
| **Europe** | | | | | |
| Albania | Yes | **Indication**: 12+ with immunosuppression | Yes | **Indication**: 60+; people with underlying conditions; frontline workers | https://albaniandailynews.com/news/albania-to-start-covid-19-booster-shots-for-60-plus-citizens |
| Austria | Yes | **Indication**: Personnel in old people's, nursing and retirement homes; health care personnel; personnel in mobile care, support, nursing and 24-hour care as well as caring relatives; staff in educational institutions (childcare, school, university, etc.)<br>**Interval**: 270-365 d | Yes | **Indication**: 16+; 12+ at risk<br>**Interval**: 270-365 d; 180-270 d | https://www.thelocal.at/20210827/first-austrian-state-to-start-covid-booster-shots-from-monday/ |
| Azerbaijan | No data | | Yes | **Indication**: HCW; 60+ | https://www.azernews.az/nation/184187.html |
| Belgium | Yes | **Indication**: People with diminished immunity, caused by a particular disease or by treatment | Yes | **Indication**: Residents of nursing homes; 65+; patients in assisted living apartments; people in day care centers, psychogeriatric facilities and | https://www.health.belgium.be/fr/news/conference-interministerielle-sante-publique-0<br>https://www.info-coronavirus.be/en/vaccination/#faq |

| | | | | | |
|---|---|---|---|---|---|
| | | | | psychiatric care homes | |
| Bulgaria | Yes | **Indication**: people with immunosuppression<br>**Interval**: 28 d | Yes | **Indication**: Persons in old people's homes and social institutions for residential accommodation; HCW; 65+<br>**Interval**: 180 d | https://coronavirus.bg/bg/news/251 |
| Cyprus | No data | | Yes | **Indication:** 60+; People with underlying condition<br>**Interval**: 180 d | https://www.pio.gov.cy/coronavirus/uploads/18102021--%CE%A7%CE%BF%CF%81%CE%AE%CE%B3%CE%B7%CF%83%CE%B7%203%CE%B7%CF%82%20%CE%B4%CF%8C%CF%83%CE%B7%CF%82%20%CE%B5%CE%BC%CE%B2%CE%BF%CE%BB%CE%AF%CE%BF%CF%85%20%CF%83%CE%B5%20%CE%AC%CF%84%CE%BF%CE%BC%CE%B1%2060%20%CE%B5%CF%84%CF%8E%CE%BD%20%CE%BA%CE%B1%CE%B9%20%CE%AC%CE%BD%CF%89.pdf |
| Czechia | Yes | **Indication**: People with moderate to severe immune suppression | Yes | **Indication**: 12+<br>**Interval**: 180 d | http://www.news.cn/english/2021-08/31/c_1310158121.htm<br>https://covid.gov.cz/en/situations/register-vaccination/booster-and-additional-dose |
| Denmark | Yes | **Indication**: People with severely impaired immune system<br>**Interval**: 30-270 d | Yes | **Indication**: 65+; essential workers<br>**Interval**: 180 d | https://www.sst.dk/da/Nyheder/2021/Personer-med-et-svaert-nedsat-immunforsvar-bliver-nu-tilbudt-en-3_-dosis-COVID-19-vaccine<br>https://www.sst.dk/da/corona/Vaccination/Revaccination-af-ud |

| | | | | | |
|---|---|---|---|---|---|
| | | | | | valgte-grupper |
| Estonia | Yes | **Indication:** People with a weakened immune system | Yes | **Indication**: 65+; 18+ requiring for a booster dose<br>**Interval**: 180 d; 240 d | https://vaktsineeri.ee/en/news/third-vaccine-doses-may-be-available-in-october/<br>https://vaktsineeri.ee/en/news/booster-doses-will-become-available-for-key-workers/(65+,care-home inhabitant) |
| Finland | Yes | **Indication**: People with severe immunosuppression; people receiving the first and second dose less than six weeks apart | Yes | **Indication**: HCW; elderly residents of nursing home and the employees caring for them<br>**Interval**: 21-28 d | https://thl.fi/en/web/thlfi-en/-/thl-proposes-a-third-coronavirus-vaccine-dose-for-limited-groups |
| France | No data | | Yes | **Indication**: 65+; certain particularly fragile patients; people who have been vaccinated with the Janssen vaccine; Health and medico-social professionals; medical transport professionals; those 18 and over who are immunocompromised<br>**Interval**: 180 d | https://vaccination-info-service.fr/Les-maladies-et-leurs-vaccins/COVID-19 |
| Germany | Yes | **Indication**: People who may | No data | **Indication**: 70+; old people in | https://www.bundesgesundheitsministerium.de/coronavirus/fa |

| | | | | | |
|---|---|---|---|---|---|
| | | not have a sufficient or rapidly decreasing immune response after a complete vaccination, including residents of care facilities, facilities for people with disabilities, other facilities with vulnerable groups, people with immunodeficiency or immunosuppression **Interval**: 180 d | | nursing home; HCW **Interval**: 180 d | q-covid-19-impfung.html#c21988 https://www.rki.de/DE/Content/Kommissionen/STIKO/Empfehlungen/PM_2021-10-07.html;jsessionid=A597C1109F012BB859E304E5DFDAF898.internet101 |
| Greece | No data | | Yes | **Indication**: 60+; HCW | https://www.euractiv.com/section/politics/short_news/greece-offers-booster-dose-as-thousands-of-health-workers-suspended/ |
| Hungary | No data | | Yes | **Indication**: 18+ **Interval**: 120 d | https://koronavirus.gov.hu/cikkek/magyarorszag-az-elso-ahol-mar-harmadik-oltasra-lehet-idopontot-foglalni |
| Iceland | No data | | Yes | **Indication**: 12+ with a history of COVID-19; individuals without history of COVID-19 antibodies who were vaccinated with the Janssen vaccine; nursing home residents and other very vulnerable welfare service | https://www.landlaeknir.is/um-embaettid/greinar/grein/item47460/booster-vaccinations-for-covid-19- |

| | | | | | |
|---|---|---|---|---|---|
| | | | | recipients (60 years and older only); highly immunosuppressed individuals with continuing immunosuppression; 60+ outside nursing homes; frontline HCW | |
| Ireland | Yes | **Indication**: People with cancer, kidney diseases, HIV, transplants, genetic diseases, High dose systemic steroids, immunocompromise; people with following treatment in the last 6 months: Cyclophosphamide, Rituximab, Alemtuzumab, Cladribine, Ocrelizumab<br>**Interval**: 60 d | Yes | **Indication**: 65+ living in Long Term Residential Care Facilities; 80+ living in the community | https://www2.hse.ie/screening-and-vaccinations/covid-19-vaccine/get-the-vaccine/weak-immune-system/<br>https://www2.hse.ie/screening-and-vaccinations/covid-19-vaccine/get-the-vaccine/covid-19-vaccine-booster-dose/ |
| Israel | No data | | Yes | **Indication**: 12+<br>**Interval**: 150 d | https://govextra.gov.il/ministry-of-health/covid19-vaccine/en-covid19-vaccination-information/<br>https://govextra.gov.il/ministry-of-health/covid19-vaccine/en-covid19-vaccine-faqs/ |
| Italy | Yes | **Indication**: people who | Yes | **Indication**: 60+; vulnerable | https://www.aifa.gov.it/en/domande-e-risposte-su-vaccini-covi |

| | | | | | |
|---|---|---|---|---|---|
| | | received a solid organ transplant or who are immunocompromised<br>**Interval**: 28 d | | people | d-19<br>https://www.salute.gov.it/portale/nuovocoronavirus/dettaglioComunicatiNuovoCoronavirus.jsp?lingua=italiano&id=5835 |
| Kazakhstan | No data | | Yes | **Indication**: 12+<br>**Interval**: 180-270 d | https://primeminister.kz/en/news/reviews/deni-sau-ult-kazakstannyn-densaulyk-saktau-ministrliginin-ulttyk-zhobasy-kanday-bagyttardy-iske-asyrudy-kamtidy-1895549 |
| Kyrgyzstan | No policy | | No policy | | |
| Latvia | Yes | **Indication**: People with high immunosuppression<br>**Interval**: 28 d | No data | **Indication**: 65+; 18+ in nursing home; HCW<br>**Interval**: 180 d | https://www.vm.gov.lv/lv/jaunums/ari-latvija-saks-treso-vakcinu-devu-pret-covid-19-administresanu-ka-pirmie-tas-sanems-cilveki-ar-butiski-novajinatu-imuno-sistemu<br>https://www.vm.gov.lv/lv/jaunums/uzsak-papilddevas-3-devas-vakcinaciju-pret-covid-19-senioriem-65-veselibas-aprupes-sistemas-darbiniekiem-un-sac-klientiem |
| Lithuania | Yes | **Indication**: High-risk patients with onco-hematological diseases, receiving treatment for onco-hematological diseases, on dialysis or after organ transplantation, with autoimmune diseases when receiving immunosuppressive | Yes | **Indication**: HCW; people with underlying conditions; 65+<br>**Interval**: 180 d | https://sam.lrv.lt/lt/naujienos/nustatyta-tvarka-kaip-trecia-doze-bus-revakcinuojami-imunosupresiniai-pacientai<br>https://koronastop.lrv.lt/lt/duk/vakcinacija-nuo-covid-19/revakcinacija#item-1054 |

| | | | | | |
|---|---|---|---|---|---|
| | | therapy | | | |
| Luxembourg | Yes | **Indication**: Immunocompromised patients | Yes | **Indication**: 75+; older adults in nursing home; people on dialysis<br>**Interval**: 180 d | https://sante.public.lu/fr/espace-professionnel/recommandations/conseil-maladies-infectieuses/covid-19/covid-19-annexes/CSMI-recommandation-3eme-dose-vaccin-COVID-19-personnes-immunodeprimees.pdf<br>https://msan.gouvernement.lu/en/actualites.gouvernement%2Ben%2Bactualites%2Btoutes_actualites%2Bcommuniques%2B2021%2B09-septembre%2B14-vaccin-dose-additionnelle.html |
| Malta | Yes | **Indication**: Old frail people; people with immunosuppression<br>**Interval**: 21 d | No data | | https://deputyprimeminister.gov.mt/en/health-promotion/covid-19/Pages/vaccines.aspx<br>https://www.independent.com.mt/articles/2021-09-13/local-news/Malta-starts-giving-Covid-19-booster-shots-to-immunosuppressed-patients-6736236685 |
| Montenegro | Yes | **Indication**: Vulnerable people<br>**Interval**: 60 d | No data | | https://www.covidodgovor.me/me/cesta-pitanja |
| Netherlands | Yes | **Indication**: People with immunosuppression | Under discussion | | https://www.rivm.nl/en/covid-19-vaccination/vaccines/immunocompromised-patients#rm-why-are-some-people-with-severely-impaired-immunity-receiving-a-third-vaccination-620821-more |
| North Macedonia | No data | | Yes | Indication: 60+; HCW; people with immunosuppression<br>Interval: 180d | https://koronavirus.gov.mk/vesti/223104 |

| | | | | | |
|---|---|---|---|---|---|
| Norway | Yes | **Indication**: People with immunosuppression | Yes | **Indication**: 65+<br>**Interval**: 180 d | https://www.fhi.no/publ/brev/ytterligere-informasjon-om-en-tredje-dose-til-de-med-alvorlig-nedsatt-immun/<br>https://www.fhi.no/en/news/2021/booster-dose-for-the-elderly-and-nursing-home-residents/ |
| Poland | Yes | **Indication**: 12+ with immunosuppression<br>**Interval**: At least 28 d | Yes | **Indication**: 50+; HCW<br>**Interval**: 180 d | https://www.gov.pl/web/zdrowie/komunikat-nr-12-ministra-zdrowia-w-sprawie-szczepien-przeciw-covid-19-dawka-przypominajaca-oraz-dawka-dodatkowa-uzupelniajaca-schemat-podstawowy |
| Portugal | Yes | **Indication**: People with immunosuppression | Yes | **Indication**: 65+; people in nursing home<br>**Interval**: 180 d | https://covid19.min-saude.pt/terceira-dose-comeca-a-ser-administrada-a-idosos-com-mais-de-65-anos-e-residentes-em-lares/ |
| Romania | No data | | Yes | **Indication**: 12+<br>**Interval**: 180 d | https://balkaninsight.com/2021/09/23/romania-to-offer-vaccine-booster-doses-as-covid-19-surges/ |
| Russia | No data | | Yes | **Indication**: 18+<br>**Interval**: 180 d | |
| Serbia | No data | | Yes | **Indication**: 18+<br>**Interval**: 180 d | https://www.srbija.gov.rs/vest/en/177106/third-dose-of-covid-19-vaccine-available-as-of-tomorrow.php |
| Slovakia | No data | | Yes | **Indication**: 12+<br>**Interval**: 240 d | https://worldakkam.com/covid-booster-shots-will-be-available-in-slovakia-starting-monday/310423/ |
| Slovenia | Yes | **Indication**: 70+ and particularly vulnerable chronic patients | No data | | https://www.cepimose.si/cepljenje-proti-covidu-19/pogosta-vprasanja-in-odgovori/ |

| | | | | | |
|---|---|---|---|---|---|
| Spain | Yes | **Indication**: People with immunosuppression; 40+ with Down Syndrome<br>**Interval**: 28 d | Yes | **Indication**: 70+; inmates in nursing homes<br>**Interval**: 180 d | https://www.vacunacovid.gob.es/preguntas-y-respuestas/quienes-recibiran-una-dosis-adicional-de-la-vacuna<br>https://www.vacunacovid.gob.es/preguntas-y-respuestas |
| Sweden | Yes | **Indication**: 18+ with severe immunosuppression<br>**Interval**: 56 d | Yes | **Indication**: People in nursing home or receiving home care; 80+<br>**Interval**: 180 d | https://www.folkhalsomyndigheten.se/smittskydd-beredskap/utbrott/aktuella-utbrott/covid-19/vaccination-mot-covid-19/fragor-och-svar-om-vaccination-mot-covid-19/<br>https://www.1177.se/en/other-languages/other-languages/covid-19/vaccin-engelska/#section-133829 |
| Switzerland | Yes | **Indication**: people with severe immunosuppression | Under discussion | | https://www.ge.ch/en/getting-vaccinated-against-covid-19/faq-vaccination-against-covid-19 |
| Turkey | Yes | | No data | **Indication**: HCW at high risk; 50+<br>**Interval**: 90 d | https://covid19asi.saglik.gov.tr/EN-78316/frequently-asked-questions.html?Sayfa=1 |
| United Kingdom | Yes | **Indication**: People with immunosuppression<br>**Interval**: 60 d | Yes | **Indication**: 50+; HCW; 16-49 at risk of severe illness; pregnant women<br>**Interval**: 180 d | https://www.gov.uk/government/news/jcvi-issues-updated-advice-on-covid-19-booster-vaccination<br>https://www.nhs.uk/conditions/coronavirus-covid-19/coronavirus-vaccination/coronavirus-booster-vaccine/ |
| **South-East Asia** | | | | | |
| India | No data | | Under discussion | | |
| Indonesia | No data | | Yes | **Indication**: HCW | https://www.kemkes.go.id/article/view/21080200001/kemenke |

| | | | | | |
|---|---|---|---|---|---|
| | | | | | s-tegaskan-vaksinasi-booster-hanya-untuk-tenaga-kesehatan.html |
| Nepal | No data | | No policy | | |
| New Zealand | Yes | **Indication**: people with severe immunosuppression<br>**Interval**: 56 d | Under discussion | | https://www.health.govt.nz/our-work/diseases-and-conditions/covid-19-novel-coronavirus/covid-19-vaccines/covid-19-vaccine-health-advice/covid-19-vaccine-severely-immunocompromised-people |
| Sri Lanka | No data | | Yes | **Indication**: Frontline workers; tourism staff | https://www.reuters.com/world/asia-pacific/sri-lanka-orders-covid-19-booster-shots-frontline-workers-tourism-industry-2021-10-22/ |
| Thailand | No data | | Yes | **Indication**: 12+ | |
| Bhutan | No data | | Under discussion | | |
| **Western Pacific** | | | | | |
| Australia | Yes | **Indication**: 12+ with immunosuppression<br>**Interval**: 60-180 d | No policy | | https://www.health.gov.au/ministers/the-hon-greg-hunt-mp/media/booster-shot-for-severely-immunocompromised-australians |
| Cambodia | No data | | Yes | **Indication**: 18+<br>**Interval**: 120 d | https://cambodianewsservice.com/cambodia-to-administer-booster-shots-to-general-public-from-mid-october/ |
| China | No data | | Yes | **Indication:** People at high risk of exposure; 60+; international travelers | http://kw.beijing.gov.cn/art/2021/9/26/art_6722_613586.html |

| | | | | | |
|---|---|---|---|---|---|
| Cook Islands | No policy | | No policy | | |
| Japan | Yes | **Interval**: 240 d | Under discussion | | https://www.mhlw.go.jp/content/10601000/000833964.pd |
| Laos | No policy | | No policy | | |
| Malaysia | No data | | No data | **Indication**: 18+<br>**Interval**: 180 d | https://covid-19.moh.gov.my/semasa-kkm/2021/10/kelulusan-bersyarat-pemberian-dos-penggalak-booster-dose |
| Micronesia | No data | | Yes | **Indication**: 65+; 18+ with underlying conditions; 18+ at high risk of SARS-CoV-2 exposure | https://ghs.guam.gov/jic-release-no-811-four-covid-19-related-fatalities-reported-covid-19-updates-60-hospitalized-165 |
| Mongolia | No data | | Yes | | https://akipress.com/news:663617:Mongolia_vaccinates_359,447_people_with_3rd_dose_of_coronavirus_vaccine/ |
| Palau | Yes | **Indication**: people with immunosuppression | No data | | http://www.palauhealth.org/2019nCoV/PR-old/MHHS%20PSA%20VACCINATION%203RD%20DOSE%2009092021.pdf |
| Philippines | No data | | Under discussion | | |
| Singapore | Yes | **Indication**: People with immunosuppression; people receiving Sinovac | Yes | **Indication**: HCW; 18+ in institutionalised settings; 30+<br>**Interval**: 180d | https://www.moh.gov.sg/news-highlights/details/updates-to-healthcare-protocols-and-implementation-of-vaccine-booster-strategy_10Sep2021<br>https://www.gov.sg/article/expanding-vaccine-booster-programme-to-more-individuals |

| | | | | | |
|---|---|---|---|---|---|
| South Korea | No data | | Yes | **Indication**: 75+ | https://www.reuters.com/world/asia-pacific/skorea-vaccinate-12-17-year-olds-give-boosters-elderly-2021-09-27/ |
| Tuvalu | No policy | | No policy | | |
| Vanuatu | No policy | | No policy | | |

Abbreviation: d, days.

**Table S4. Country lists of selling/donating or receiving COVID-19 vaccines**

| Country role | Country lists |
|---|---|
| Selling or donating countries | Australia; Bahrain; Bhutan; Canada; China; Colombia; Iceland; Japan; Korea; Kuwait; Mauritius; Mexico; Monaco; New Zealand; Norway; Oman; The Philippines; Qatar; Saudi Arabia; Singapore; Switzerland; Austria; Belgium; Croatia; Denmark; Estonia; Finland; France; Germany; Greece; Ireland; Italy; Luxembourg; Malta; Netherlands; Poland; Portugal; Spain; Sweden; United Kingdom; United States; Vietnam; United Arab Emirates. |
| Recipient countries | Afghanistan; Albania; Algeria; Andorra; Angola; Antigua and Barbuda; Argentina; Armenia; Australia; Azerbaijan; Bahamas; Bahrain; Bangladesh; Barbados; Belize; Benin; Bhutan; Bolivia; Bosnia and Herzegovina; Botswana; Brazil; Brunei Darussalam; Burkina Faso; Cabo Verde; Cambodia; Cameroon; Canada; Central African Republic; Chad; Chile; China; Colombia; Comoros; Congo, Dem. Rep.; Congo, Rep.; Costa Rica; Cote d'Ivoire; Djibouti; Dominica; Dominican Republic; Ecuador; Egypt, Arab Rep.; El Salvador; Eswatini; Ethiopia; Fiji; Gabon; Gambia, The; Georgia; Ghana; Grenada; Guatemala; Guinea; Guinea-Bissau; Guyana; Haiti; Honduras; India4; Indonesia; Iran; Iraq; Israel; Jamaica; Jordan; Kenya; Kiribati; Korea, Dem. People's Rep.; Kosovo; Kuwait; Kyrgyz Republic; Lao PDR; Lebanon; Lesotho; Liberia; Libya; Madagascar; Malawi; Malaysia; Maldives; Mali; Mauritania; Mauritius; Mexico; Micronesia; Moldova; Monaco; Mongolia; Montenegro; Morocco; Mozambique; Myanmar; Namibia; Nauru; Nepal; New Zealand; Nicaragua; Niger; Nigeria; North Korea; North Macedonia; Oman; Pakistan; Panama; Papua New Guinea; Paraguay; Peru; Philippines; Qatar; Rwanda; Samoa; Sao Tome and Principe; Saudi Arabia; Senegal; Serbia; Sierra Leone; Singapore; Solomon Islands; Somalia; South Africa; South Sudan; South Korea; Sri Lanka; St. Kitts and Nevis; St. Lucia; St. Vincent and the Grenadines; Sudan; Suriname; Syria; Tajikistan; Timor-Leste; Togo; Tonga; Trinidad and Tobago; Tunisia; Tuvalu; Uganda; Ukraine; United Arab Emirates; United Kingdom; and Northern Ireland; Tanzania; Uruguay; Uzbekistan; Vanuatu; Venezuela; Vietnam; West Bank and Gaza; Yemen, Rep.; Zambia; Zimbabwe. |

The data are derived from COVAX[2].

### Target population for primary vaccination

#### *Method to estimate the size of target population*

##### Metrices

Some metrices have been collected to estimate target population: 1) **Total population estimates**. The age-specific population estimates by country in 2020 were mainly derived from the UN World Population Prospects, supplemented by the WorldPop datasets for some countries with age-specific proportion unavailable. 2) **The indication groups for primary vaccination**. The population groups who are eligible for a COVID-19 vaccine, recommended by governments/health departments. Indication groups varied by countries, which might be general population with a certain age group or some specific population groups, e.g., frontline workers. 3) **The contraindication groups**. The population who should not get vaccinated against COVD-19 recommended by governments/health departments, e.g., pregnant women, people with certain underlying conditions (i.e., bleeding disorders and immune suppression), and those previously infected with SARS-CoV-2. They should be excluded from the indication groups according to the vaccination policy in each country.

##### Definition of special population groups

Two underlying conditions are considered in this study: bleeding disorders and immune suppression. A bleeding disorder is defined as condition in which the blood's ability to clot is impaired or at thrombotic states[3]. Diseases that would cause bleeding disorder are considered as a bleeding problem. In the case of immune suppression, we include categories of cancers with direct immune suppression and cancers with possible immune suppression (from treatment therapy), as well as HIV/AIDS without receiving antiretroviral therapy (ART) based on Clark's method[4]. The categories and definitions of special population groups are presented in Supplementary Table 5, in which specific causes of bleeding disorders and immune suppression defined by GBD study group were listed.

**Table S5. Categories and definitions of special population groups belonging to indication and contraindication lists**

| **Population groups** | **Definition** | **Size data avail able** | **Data source** |
|---|---|---|---|
| **Pregnancy-related people** | | | |
| Pregnant women | - | Yes | Wang W et al, BMJ[5] |
| Breastfeeding (lactation) women | - | No | - |
| Plan to get pregnancy | - | No | - |
| **People working at potentially greater risk of SARS-CoV-2 infection** | | | |
| Healthcare workers | Including healthcare workers, nurses and midwives | Yes | Wang W et al, BMJ[5] |
| People maintaining society safety and national security | Police and military | Yes | Wang W et al, BMJ[5] |

| | | | |
|---|---|---|---|
| Other essential workers | People engaging in electricity, gas, water, food, steam, air conditioning, and accommodation supply; participating in sewerage, waste management, and remediation activities; involved in domestic transportation and storage | Yes | Wang W et al, BMJ[5] |

| | | | |
|---|---|---|---|
| **People at higher risk of severe COVID-19 disease** | | | |
| Residents living in nursing homes/long-care homes/centers/facilities/institutions/settings or receiving home care | Elderly people receiving long-term care at a residential care facility and at home | Partially | https://www.who.int/data/maternal-newborn-child-adolescent-ageing/indicator-explorer-new/mca/percentage-of-older-people-receiving-long-term-care-at-a-residential-care-facility-and-at-home |
| Vulnerable people with high risk of experiencing irreversible and devastating harm from COVID-19 due to their health conditions, which included the elderly and people with underlying medical conditions. | Underlying conditions included cardiovascular disease, chronic kidney disease, chronic respiratory disease, chronic | Yes | Clark A et al. Lancet GH[4]<br>Wang W et al, BMJ[5] |

| | |
|---|---|
| | liver disease, diabetes, cancer with direct immunosuppression, cancer without direct immunosuppression but with possible immunosuppression caused by treatment, HIV or AIDS, tuberculosis (excluding latent |

| | |
|---|---|
| | infections), chronic neurological disorders, and sickle cell disorders. And target population in this category was classified into (1) people younger than 60 years with at least one underlying condition; (2) people aged 60 |

| | | | |
|---|---|---|---|
| | years or older with at least one underlying conditions; (3) people aged 80 years or older without any underlying conditions. | | |
| People with (moderate to severe) immunosuppression | **Cancers with direct immune suppression:** Hodgkin lymphoma; Non-Hodgkin lymphoma; Multiple myeloma; | Yes | Estimated used data from GBD 2019[6] |

| | |
|---|---|
| | Acute lymphoid leukemia; Chronic lymphoid leukemia; Acute myeloid leukemia; Chronic myeloid leukemia; Other leukemia; Other malignant neoplasms; Myelodysplastic, myeloproliferative, and other hematopoie |

| | |
|---|---|
| | tic neoplasms **Cancers with possible immune suppression (from treatment therapy):** Lip and oral cavity cancer; Nasopharynx cancer; Other pharynx cancer; Esophageal cancer; Stomach cancer; Colon and |

| | |
|---|---|
| | rectum cancer; Liver cancer due to hepatitis B; Liver cancer due to hepatitis B; Liver cancer due to hepatitis B; Liver cancer due to hepatitis C; Liver cancer due to alcohol use; Liver cancer due to NASH; Liver cancer due to other |

| | |
|---|---|
| | causes; Gallbladder and biliary tract cancer; Pancreatic cancer; Larynx cancer; Tracheal, bronchus, and lung cancer; Malignant skin melanoma; Breast cancer; Cervical cancer; Uterine cancer; Ovarian cancer; |

| | | | |
|---|---|---|---|
| | Prostate cancer; Testicular cancer; Kidney cancer; Bladder cancer; Brain and nervous system cancer; Thyroid cancer; Mesothelioma **HIV/AIDS without receiving ART**** | | |
| People with bleeding disorders | Other nutritional deficiencies | Yes | Estimated used data from GBD 2019[6] |

| | |
|---|---|
| | ; Acute hepatitis A; Acute hepatitis B; Acute hepatitis C; Acute hepatitis E; Liver cancer due to hepatitis B; Liver cancer due to hepatitis C; Liver cancer due to alcohol use; Liver cancer due to other causes; Liver cancer due |

| | |
|---|---|
| | to NASH; Cirrhosis and other chronic liver diseases due to hepatitis B; Cirrhosis and other chronic liver diseases due to hepatitis C; Cirrhosis and other chronic liver diseases due to alcohol use; Cirrhosis |

| | |
|---|---|
| | and other chronic liver diseases due to other causes; Cirrhosis and other chronic liver diseases due to NAFLD; Endocrine, metabolic, blood, and immune disorders; Dengue; Yellow fever; Multiple myeloma; |

| | |
|---|---|
| | Acute lymphoid leukemia; Chronic lymphoid leukemia; Acute myeloid leukemia; Chronic myeloid leukemia; Other leukemia; Myelodysplastic, myeloproliferative, and other hematopoietic neoplasms; Venomous |

| | | | |
|---|---|---|---|
| | animal contact | | |
| People on dialysis | - | No | - |
| **Others** | | | |
| International travelers | - | No | - |
| People previously infected with SARS-CoV-2 | COVID-19 cumulative cases | Yes | |
| People receiving vaccines from Janssen & Janssen, Sinovac or Sinopharm | - | Partially, and was available in Chile and Uruguay* | |

* For those data that are not available, we excluded them in the calculation.

** ART coverage among people living with HIV is derived from WHO[7] and published literature[8,9]. We assumed that countries with such value unavailable are replaced with regional data. For example, we replaced ART coverage in Sweden with average value in Europe Region (71.0%).

**Estimate peoples of population size with bleeding disorders and immune suppression**

Since the prevalence of the sub-categories of each disease (bleeding disorders or immune suppression) was separately reported by GBD, one person can simultaneously suffer more than one sub-category of such disease, namely multimobidity. Thus, we estimated the number of people who suffered at least one bleeding disorders- or immune suppression-related conditions, rather than directly adding the number of sub-categories of bleeding disorders or immune suppression.

We mainly adopted the Clark's method[4] to estimate the size of individuals with bleeding disorders or immune suppression. Here, we briefly explain this process. First, data on the prevalence (***p***) of sub-categories of bleeding disorders or immune suppression were extracted by age and country from GBD 2019[6]. Then, the expected proportion of individuals with at least one bleeding problems or at least one immune suppression problems were estimated, which we refer to ***e*** here. $\mathbf{e}_{\text{bleeding}}$ for bleeding problems was acquired by 1 minus the probability of not having a condition in any of the 26 bleeding problems $c_{\text{bleeding i}}$: $1 - [1 - p(c_{\text{bleeding1}})]\times[1 - p(c_{\text{bleeding2}})]\times\ldots\times[1 - p(c_{\text{bleeding27}})]$; and $\mathbf{e}_{\text{immuno}}$ for immunosuppression was acquired by 1 minus the probability of not having a condition in any of the 40 immunosuppression problems $c_{\text{immunoi}}$: $1 - [1 - p(c_{\text{immuno1}})]\times[1 - p(c_{\text{immuno2}})]\times\ldots\times[1 - p(c_{\text{immuno40}})]$. Subsequently, the observed proportion (P) of people with at least one underlying condition was calculated by $P = \boldsymbol{e} \times \mathbf{r}$, where ***r*** was the ratio between the observed and expected percentage of individuals with at least one condition, with details in Clark et al[4].

14 types of sub-diseases are considered as both bleeding disorders and immune suppression, including multiple myeloma, acute lymphoid leukemia, chronic lymphoid leukemia, acute myeloid leukemia, chronic myeloid leukemia, other leukemia, myelodysplastic, myeloproliferative, other hematopoietic neoplasms, liver cancer due to hepatitis B, liver cancer due to hepatitis C, liver cancer due to alcohol use, liver cancer due to other causes, and liver cancer due to nonalcoholic steatohepatitis (NASH). When calculating the proportion of individuals with at least one underlying conditions for these sub-diseases, we counted them once: $(1 - [1 - p\,(c_{\text{bleeding1}})] \times [1 - p\,(c_{\text{bleeding2}})] \times\ldots\times [1 - p\,(c_{\text{bleeding26}})] \times [1 - p\,(c_{\text{immuno1}})] \times [1 - p\,(c_{\text{immuno2}})] \times\ldots\times [1 - p\,(c_{\text{immuno26}})]) \times r$.

**Estimate population size of target population**

We calculated the country-specific number of contraindication groups by adding the size of pregnant women, people with certain underlying conditions, and those previously infected with SARS-CoV-2 according to the national immunization policy. Then, we estimated the size of target population for primary immunization by subtracting the population of contraindications group from the population of indication groups.

**Table S6. Global, regional, and national target population.**

| Locations | No. of total population (million) | No. of unapproved age groups (million) | No. of contraindication population (million) | Target population (million) |
|---|---|---|---|---|
| Global | | | | |
| Total | 7750.0 | 2074.5 | 132.1 | 5525.6 |
| WHO regions | | | | |
| AFR | 1120.2 | 587.7 | 9.4 | 531.1 |
| AMR | 1018.1 | 186.9 | 40.2 | 791.1 |
| EMR | 725.7 | 240.9 | 8.7 | 476.1 |
| EUR | 932.9 | 149.2 | 23.2 | 760.4 |
| SEAR | 2021.4 | 618.2 | 29.9 | 1347.5 |
| WPR | 1931.7 | 291.6 | 20.7 | 1619.5 |
| Country | | | | |
| Afghanistan | 38.9 | 19.1 | - | 19.8 |
| Albania | 2.9 | 0.6 | - | 2.3 |
| Algeria | 43.9 | 15.3 | - | 28.6 |
| Andorra | 0.1 | 0.0 | - | 0.1 |
| Angola | 32.9 | 17.5 | - | 15.4 |
| Antigua and Barbuda | 0.1 | 0.0 | 0.0 | 0.1 |
| Argentina | 45.2 | 8.9 | 0.8 | 35.5 |
| Armenia | 3.0 | 0.7 | 0.0 | 2.2 |
| Australia | 25.5 | 4.0 | 0.3 | 21.2 |
| Austria | 9.0 | 1.0 | 0.1 | 7.9 |
| Azerbaijan | 10.1 | 2.0 | 0.2 | 8.0 |
| Bahamas | 0.4 | 0.1 | - | 0.3 |
| Bahrain | 1.7 | 0.3 | 0.0 | 1.4 |
| Bangladesh | 164.7 | 53.4 | - | 111.3 |
| Barbados | 0.3 | 0.0 | - | 0.2 |
| Belarus | 9.4 | 1.7 | 0.3 | 7.4 |
| Belgium | 11.6 | 1.6 | 0.1 | 9.9 |
| Belize | 0.4 | 0.1 | - | 0.3 |
| Benin | 12.1 | 5.9 | - | 6.2 |
| Bhutan | 0.8 | 0.2 | 0.0 | 0.6 |
| Bolivia | 11.7 | 3.8 | 0.3 | 7.7 |
| Bosnia and Herzegovina | 3.3 | 0.4 | - | 2.9 |
| Botswana | 2.4 | 0.9 | - | 1.4 |

| | | | | |
|---|---|---|---|---|
| Brazil | 212.6 | 35.0 | 3.0 | 174.6 |
| Brunei | 0.4 | 0.1 | 0.0 | 0.4 |
| Bulgaria | 6.9 | 0.8 | - | 6.2 |
| Burkina Faso | 20.9 | 10.7 | - | 10.2 |
| Burundi | 11.9 | 11.4 | - | 1.7 |
| Cabo Verde | 0.6 | 0.2 | - | 0.4 |
| Cambodia | 16.7 | 2.1 | - | 14.6 |
| Cameroon | 26.5 | 12.9 | - | 13.6 |
| Canada | 37.7 | 4.8 | 0.4 | 32.6 |
| Central African Republic | 4.8 | 4.7 | - | 0.8 |
| Chad | 16.4 | 6.4 | - | 10.1 |
| Chile | 19.1 | 1.4 | 0.2 | 17.5 |
| China | 1439.3 | 204.8 | 15.7 | 1218.8 |
| Colombia | 50.9 | 8.9 | 0.8 | 41.2 |
| Comoros | 0.9 | 0.4 | - | 0.5 |
| Congo | 5.5 | 2.6 | - | 2.9 |
| Cook Islands | 0.0 | 0.0 | 0.0 | 0.0 |
| Costa Rica | 5.1 | 0.8 | - | 4.2 |
| Cote d'Ivoire | 26.4 | 12.7 | - | 13.6 |
| Croatia | 4.1 | 0.6 | 0.0 | 3.4 |
| Cuba | 11.3 | 0.2 | - | 11.1 |
| Cyprus | 1.2 | 0.2 | - | 1.0 |
| Czechia | 10.7 | 1.3 | 0.1 | 9.2 |
| Democratic Republic of the Congo | 89.6 | 83.4 | - | 15.5 |
| Denmark | 5.8 | 0.7 | 0.1 | 5.0 |
| Djibouti | 1.0 | 0.3 | - | 0.6 |
| Dominica | 0.1 | 0.0 | - | 0.1 |
| Dominican Republic | 10.8 | 2.4 | 0.2 | 8.2 |
| Ecuador | 17.6 | 3.9 | - | 13.7 |
| Egypt | 102.3 | 40.0 | - | 62.3 |
| El Salvador | 6.5 | 0.7 | 0.1 | 5.7 |
| Equatorial Guinea | 1.4 | 0.6 | - | 0.8 |
| Eritrea | 3.5 | - | - | - |

| | | | | |
|---|---|---|---|---|
| Estonia | 1.3 | 0.2 | 0.0 | 1.1 |
| Eswatini | 1.2 | 0.3 | - | 0.8 |
| Ethiopia | 115.0 | 87.8 | - | 27.1 |
| Fiji | 0.9 | 0.3 | 0.0 | 0.6 |
| Finland | 5.5 | 0.7 | 0.0 | 4.8 |
| France | 65.3 | 9.1 | 13.8 | 42.3 |
| Gabon | 2.2 | 0.7 | - | 1.5 |
| Gambia | 2.4 | 2.4 | - | 0.3 |
| Georgia | 4.0 | 0.9 | 0.1 | 3.0 |
| Germany | 83.8 | 9.4 | 0.8 | 73.6 |
| Ghana | 31.1 | 13.5 | - | 17.6 |
| Greece | 10.4 | 1.1 | - | 9.3 |
| Grenada | 0.1 | 0.0 | 0.0 | 0.1 |
| Guatemala | 17.9 | 7.1 | 0.4 | 10.3 |
| Guinea | 13.1 | 6.6 | - | 6.5 |
| Guinea-Bissau | 2.0 | 1.0 | - | 1.0 |
| Guyana | 0.8 | 0.3 | - | 0.5 |
| Haiti | 11.4 | 4.4 | - | 7.0 |
| Honduras | 9.9 | 3.0 | 0.2 | 6.7 |
| Hungary | 9.7 | 1.1 | 0.1 | 8.5 |
| Iceland | 0.3 | 0.1 | 0.0 | 0.3 |
| India | 1380.0 | 436.9 | 24.6 | 918.5 |
| Indonesia | 273.5 | 57.2 | 4.9 | 211.3 |
| Iran | 84.0 | 24.1 | - | 59.8 |
| Iraq | 40.2 | 17.7 | - | 22.5 |
| Ireland | 4.9 | 0.8 | 0.1 | 4.1 |
| Israel | 8.7 | 2.0 | 0.2 | 6.5 |
| Italy | 60.5 | 6.1 | 0.4 | 53.9 |
| Jamaica | 3.0 | 0.7 | 0.0 | 2.2 |
| Japan | 126.5 | 12.4 | 0.9 | 113.1 |
| Jordan | 10.2 | 2.7 | 0.2 | 7.3 |
| Kazakhstan | 18.8 | 4.6 | - | 14.2 |
| Kenya | 53.8 | 24.4 | - | 29.3 |
| Kiribati | 0.1 | 0.0 | - | 0.1 |
| Kuwait | 4.3 | 0.7 | - | 3.5 |
| Kyrgyzstan | 6.5 | 2.4 | 0.2 | 3.9 |
| Laos | 7.3 | 2.6 | 0.2 | 4.5 |

| | | | | |
|---|---|---|---|---|
| Latvia | 1.9 | 0.2 | 0.0 | 1.6 |
| Lebanon | 6.8 | 1.8 | 0.1 | 4.9 |
| Lesotho | 2.1 | 0.7 | - | 1.4 |
| Liberia | 5.1 | 2.4 | - | 2.7 |
| Libya | 6.9 | 2.3 | 0.1 | 4.5 |
| Lithuania | 2.7 | 0.4 | - | 2.4 |
| Luxembourg | 0.6 | 0.1 | - | 0.5 |
| Macedonia | 2.1 | 0.3 | - | 1.8 |
| Madagascar | 27.7 | 12.9 | - | 14.8 |
| Malawi | 19.1 | 9.6 | - | 9.6 |
| Malaysia | 32.4 | 6.1 | 0.8 | 25.5 |
| Maldives | 0.5 | 0.1 | - | 0.5 |
| Mali | 20.3 | 10.9 | - | 9.3 |
| Malta | 0.4 | 0.1 | 0.0 | 0.4 |
| Marshall Islands | 0.1 | 0.0 | - | 0.0 |
| Mauritania | 4.6 | 1.5 | - | 3.1 |
| Mauritius | 1.3 | 0.2 | - | 1.1 |
| Mexico | 128.9 | 26.6 | 29.8 | 72.5 |
| Micronesia | 0.1 | 0.0 | - | 0.1 |
| Moldova | 4.0 | 0.8 | - | 3.3 |
| Monaco | 0.0 | 0.0 | - | 0.0 |
| Mongolia | 3.3 | 0.9 | - | 2.4 |
| Montenegro | 0.6 | 0.1 | 0.0 | 0.5 |
| Morocco | 36.9 | 8.0 | 0.7 | 28.2 |
| Mozambique | 31.3 | 13.8 | 1.2 | 16.3 |
| Myanmar | 54.4 | 46.4 | - | 8.0 |
| Namibia | 2.5 | 1.1 | - | 1.5 |
| Nauru | 0.0 | 0.0 | - | 0.0 |
| Nepal | 29.1 | 10.3 | - | 18.9 |
| Netherlands | 17.1 | 2.1 | - | 15.0 |
| New Zealand | 4.8 | 0.7 | 0.1 | 4.0 |
| Nicaragua | 6.6 | 2.3 | - | 4.3 |
| Niger | 24.2 | 13.7 | - | 10.5 |
| Nigeria | 206.1 | 94.3 | 7.8 | 104.0 |
| Niue | 0.0 | 0.0 | - | 0.0 |
| North Korea | 25.8 | - | - | - |
| Norway | 5.4 | 0.7 | - | 4.7 |

| | | | | |
|---|---|---|---|---|
| Oman | 5.1 | 1.0 | - | 4.1 |
| Pakistan | 220.9 | 63.1 | 6.2 | 151.6 |
| Palau | 0.0 | 0.0 | - | 0.0 |
| Panama | 4.3 | 0.9 | - | 3.4 |
| Papua New Guinea | 8.9 | 3.7 | - | 5.2 |
| Paraguay | 7.1 | 2.5 | - | 4.7 |
| Peru | 33.0 | 9.6 | - | 23.4 |
| Philippines | 109.6 | 26.5 | 2.3 | 80.9 |
| Poland | 37.8 | 4.6 | 1.0 | 32.2 |
| Portugal | 10.2 | 1.0 | 0.1 | 9.1 |
| Qatar | 2.9 | 0.3 | 0.0 | 2.5 |
| Romania | 19.2 | 2.3 | - | 16.9 |
| Russia | 145.9 | 31.2 | 1.7 | 113.1 |
| Rwanda | 13.0 | 5.9 | 0.4 | 6.6 |
| Saint Kitts and Nevis | 0.1 | 0.0 | 0.0 | 0.0 |
| Saint Lucia | 0.2 | 0.0 | - | 0.2 |
| Saint Vincent and the Grenadines | 0.1 | 0.0 | - | 0.1 |
| Samoa | 0.2 | 0.1 | 0.0 | 0.1 |
| San Marino | 0.0 | 0.0 | 0.0 | 0.0 |
| Sao Tome and Principe | 0.2 | 0.1 | - | 0.1 |
| Saudi Arabia | 34.8 | 7.1 | 1.2 | 26.6 |
| Senegal | 16.7 | 8.2 | - | 8.5 |
| Serbia | 8.7 | 1.0 | - | 7.7 |
| Seychelles | 0.1 | 0.0 | - | 0.1 |
| Sierra Leone | 8.0 | 3.8 | - | 4.2 |
| Singapore | 5.9 | 0.6 | 0.1 | 5.2 |
| Slovakia | 5.5 | 0.7 | 0.1 | 4.7 |
| Slovenia | 2.1 | 0.3 | 0.6 | 1.3 |
| Solomon Islands | 0.7 | 0.3 | - | 0.4 |
| Somalia | 15.9 | 8.5 | - | 7.4 |
| South Africa | 59.3 | 13.8 | - | 45.5 |
| South Korea | 51.3 | 7.8 | 0.3 | 43.1 |
| South Sudan | 11.2 | 5.4 | - | 5.8 |

| | | | | |
|---|---|---|---|---|
| Spain | 46.8 | 5.2 | 0.4 | 41.2 |
| Sri Lanka | 21.4 | 4.0 | 0.3 | 17.0 |
| Sudan | 43.8 | 20.4 | - | 23.4 |
| Suriname | 0.6 | 0.2 | 0.0 | 0.4 |
| Sweden | 10.1 | 1.4 | 0.1 | 8.6 |
| Switzerland | 8.7 | 1.0 | 0.1 | 7.5 |
| Syria | 17.5 | 6.3 | - | 11.2 |
| Tajikistan | 9.5 | 4.0 | - | 5.5 |
| Tanzania | 59.7 | 30.0 | - | 29.7 |
| Thailand | 69.8 | 9.1 | - | 60.7 |
| Timor-Leste | 1.3 | 0.6 | - | 0.7 |
| Togo | 8.3 | 4.3 | - | 4.0 |
| Tonga | 0.1 | 0.0 | - | 0.1 |
| Trinidad and Tobago | 1.4 | 0.2 | 0.0 | 1.2 |
| Tunisia | 11.8 | 3.3 | - | 8.5 |
| Turkey | 84.3 | 16.1 | 1.4 | 66.9 |
| Turkmenistan | 6.0 | 2.2 | - | 3.9 |
| Tuvalu | 0.0 | 0.0 | - | 0.0 |
| Uganda | 45.7 | 17.4 | - | 28.3 |
| Ukraine | 43.7 | 5.6 | 0.4 | 37.8 |
| United Arab Emirates | 9.9 | 0.3 | 0.1 | 9.5 |
| United Kingdom | 67.9 | 9.7 | 0.7 | 57.5 |
| United States | 331.0 | 48.1 | 3.8 | 279.1 |
| Uruguay | 3.5 | 0.6 | 0.0 | 2.9 |
| Uzbekistan | 33.5 | 7.9 | - | 25.5 |
| Vanuatu | 0.3 | 0.1 | - | 0.2 |
| Venezuela | 28.4 | 9.3 | - | 19.1 |
| Vietnam | 97.3 | 18.4 | - | 79.0 |
| Yemen | 29.8 | 13.5 | - | 16.3 |
| Zambia | 18.4 | 9.4 | - | 9.0 |
| Zimbabwe | 14.9 | 5.9 | - | 9.0 |

Abbreviation: AFR, African Region; AMR, Region of Americas; EMR, Eastern Mediterranean Region; EUR, European Region; SEAR, South-East Asia Region; WPR, Western Pacific Region.

#### COVID-19 vaccine coverage

##### *Metrics included in the dataset of administered doses*

(1) **Number of vaccine doses administered**. The cumulative administered number of COVID-19 vaccine doses stratified by time, vaccine technical platform, and age group, which might include initial doses, additional doses, and booster doses.

(2) **The number of people vaccinated at least one dose**. The number of people who have received the first dose of a multiple-dose vaccine or the dose administered for a one-dose vaccine.

(3) **The number of people fully vaccinated**. The number of people who have completed their primary immunization series according to the vaccination schedule.

(4) **The number of people receiving additional/booster doses**. The number of cumulative people receiving additional or/and booster doses.

**Figure S1. Proportion administered by vaccine technical platforms**

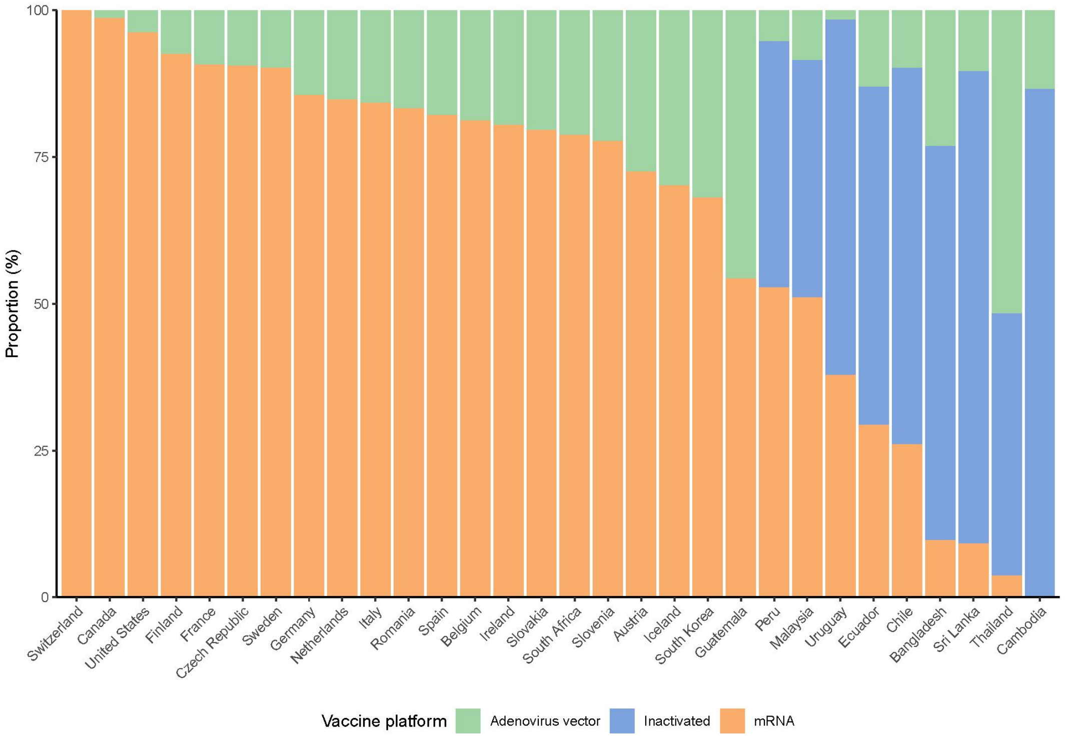

**Figure S2. Proportion administered by vaccine types.**

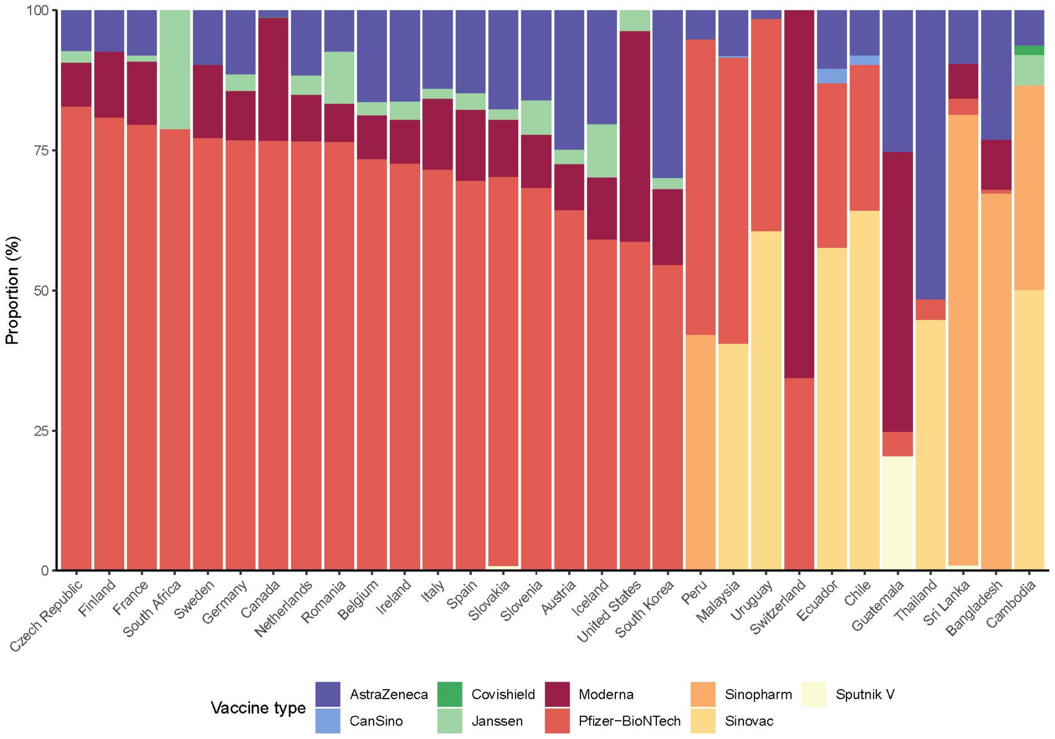

**Figure S3. Date at which achieved one dose per 100 people in total population by countries.**

The white areas represent countries that have not achieved one dose per 100 people or for which data are unavailable. The data shown here are as of October 7, 2021.

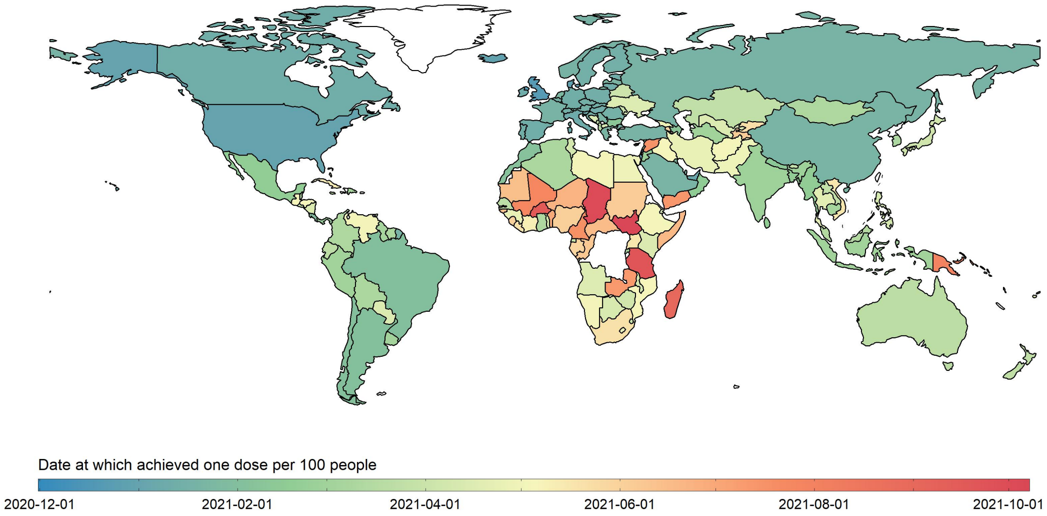

**Figure S4. Vaccine coverage over time stratified by income groups and role of vaccine seller/donor or recipient.**

Cumulative doses per 100 people by income group in total population (A) and target population (B). Cumulative doses per 100 people by role of vaccine seller/donor or recipient in total population (C) and target population (D). Income group was defined by the World Bank. The data on role of vaccine seller/donor or recipient was derived from COVAX.

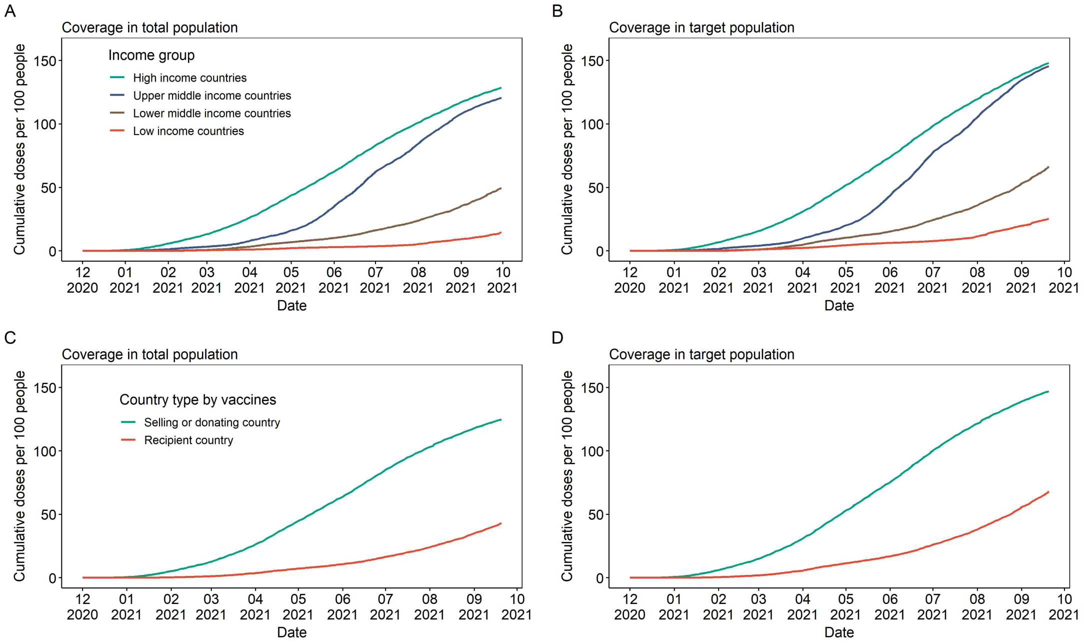

**Figure S5. Vaccine coverage stratified by SDI quintile and WHO region.**

SDI, socio-demographic index from the Institute for Health Metrics and Evaluation (IHME)[10].

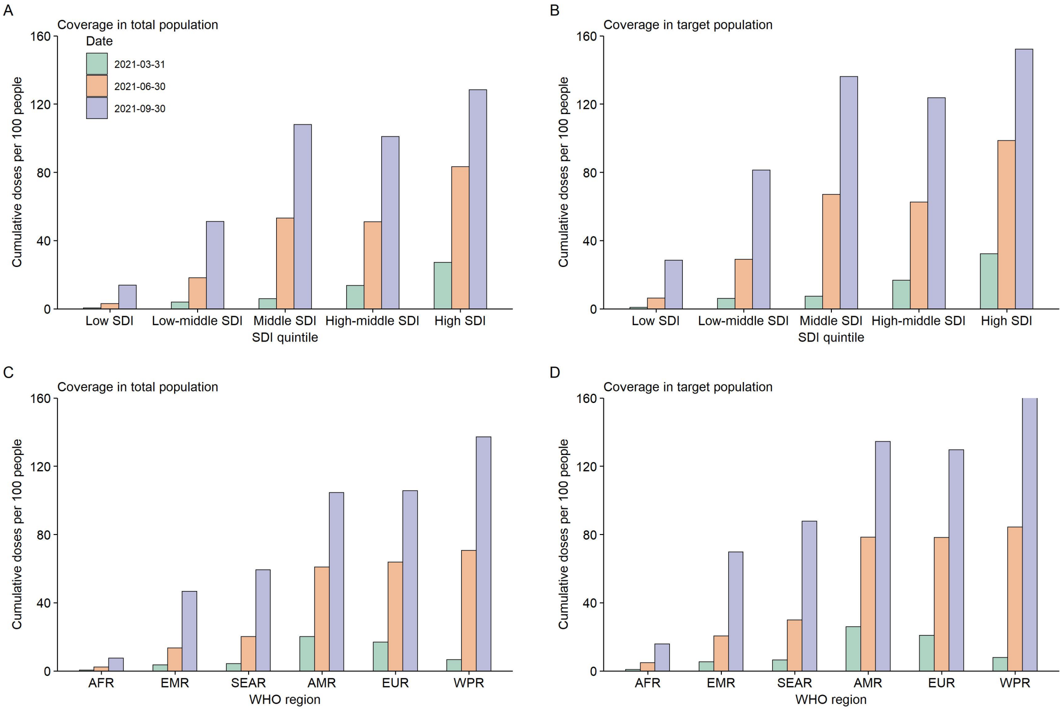

**Figure S6. Corrections between vaccine coverage and country-level vaccine acceptance.** Data on country-specific vaccine acceptance among general population was summarized through a rapid review by selecting one representative study for each country, based on comprehensive evaluation about study period, sampling method and representativeness of study participants.

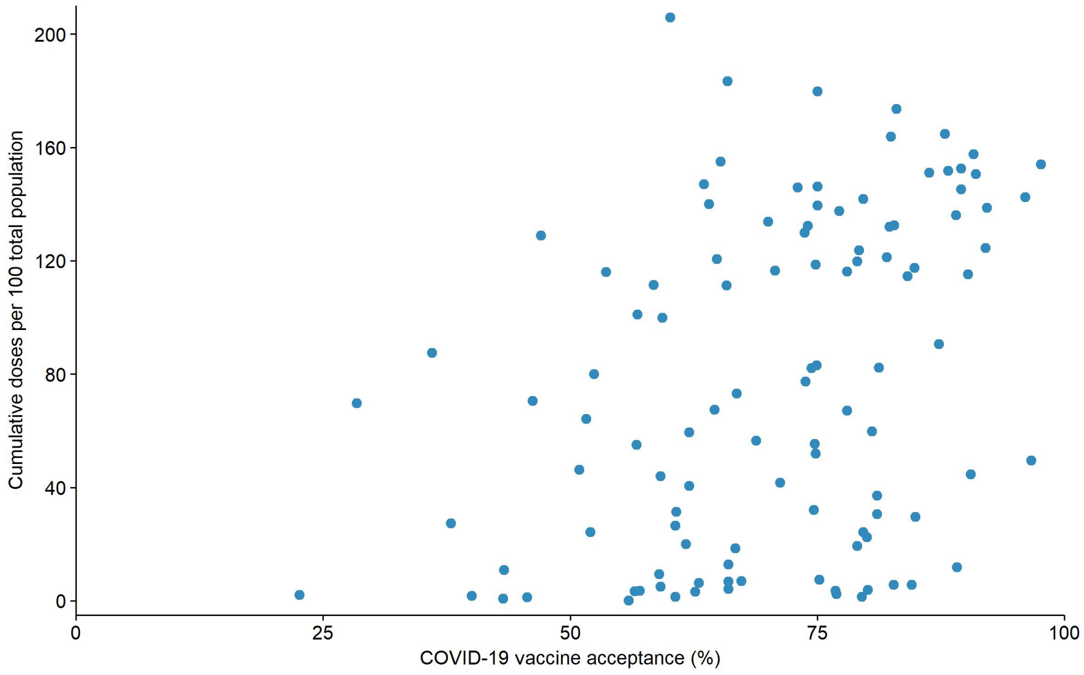

#### Demand of COVID-19 vaccine doses

##### *Method to calculate the demand of COVID-19 vaccine doses*

In primary immunization, we assumed a two-dose schedule to get the total doses required for primary immunization of target population. With an exception of previous infected COVID-19 patients, if countries recommended 1 dose for those groups, one-dose schedule was used instead. The target population for primary immunization was presented in Table S2.

In additional/booster immunization programs, we assumed a one-dose schedule for those target population recommended by each country, respectively (Table S3). Generally, the target groups for an additional dose mainly contained some specific population groups, e.g., people undergoing immunosuppression, and/or elderly, and/or specific population groups at high risk of infection (such as international travelers, front-line workers). While the target groups for a booster dose may include people with specific age, and/or people undergoing immunosuppression, and/or frontline workers, and/or people receiving Sinovac/Sinopharm vaccine.

By multiplying the number of specific target population and required doses, we obtained the doses required for primary and additional/booster immunization programs, respectively. Finally, we subtracted cumulative primary doses and additional/booster doses administered in each country to get the demand of COVID-19 vaccine doses for primary and additional/booster immunization programs, respectively (Table S7).

**Table S7. Global, regional, and national demand of vaccine dose.**

| Locations | Doses required for vaccination (millions) | | | Administered doses for vaccination (millions) | | | Current demand doses (millions) | | |
|---|---|---|---|---|---|---|---|---|---|
| | Total | Primary doses | Additional/ booster doses | Total | Primary doses | Additional/ booster doses | Total | Primary doses | Additional/ Booster doses |
| Global | | | | | | | | | |
| Total | 11956.9 | 11031.3 | 925.7 | 6427.2 | 6382.4 | 44.7 | 5537.4 | 4650.4 | 887.0 |
| WHO region | | | | | | | | | |
| AFR | 1064.6 | 1062.1 | 2.5 | 86.9 | 86.9 | - | 976.1 | 973.6 | 2.5 |
| AMR | 1760.4 | 1582.2 | 178.3 | 1094.0 | 1073.5 | 20.5 | 672.1 | 508.7 | 163.4 |
| EMR | 966.0 | 952.2 | 13.8 | 323.8 | 323.5 | 0.3 | 636.4 | 623.0 | 13.5 |
| EUR | 1854.9 | 1501.2 | 353.7 | 993.1 | 972.1 | 21.0 | 870.7 | 538.0 | 332.7 |
| SEAR | 2758.4 | 2694.9 | 63.4 | 1244.9 | 1243.3 | 1.6 | 1513.4 | 1451.6 | 61.8 |
| WPR | 3552.7 | 3238.7 | 314.0 | 2684.3 | 2683.1 | 1.3 | 868.6 | 555.6 | 313.1 |
| Countries | | | | | | | | | |
| Afghanistan | 39.6 | 39.6 | - | 2.5 | 2.5 | - | 37.1 | 37.1 | - |
| Albania | 6.0 | 4.5 | 1.5 | 1.8 | 1.8 | - | 4.2 | 2.7 | 1.5 |
| Algeria | 57.1 | 57.1 | - | 10.6 | 10.6 | - | 46.5 | 46.5 | - |
| Andorra | 0.2 | 0.1 | 0.0 | 0.1 | 0.1 | - | 0.1 | 0.0 | 0.0 |
| Angola | 30.8 | 30.8 | - | 4.2 | 4.2 | - | 26.6 | 26.6 | - |
| Antigua and Barbuda | 0.2 | 0.2 | - | 0.1 | 0.1 | - | 0.1 | 0.1 | - |
| Argentina | 71.1 | 71.1 | - | 53.6 | 53.6 | - | 17.4 | 17.4 | - |

| | | | | | | | | | |
|---|---|---|---|---|---|---|---|---|---|
| Armenia | 4.4 | 4.4 | - | 0.5 | 0.5 | - | 3.9 | 3.9 | - |
| Australia | 43.2 | 42.4 | 0.8 | 29.6 | 29.6 | - | 13.6 | 12.8 | 0.8 |
| Austria | 24.0 | 15.8 | 8.3 | 10.9 | 10.9 | - | 13.1 | 4.8 | 8.3 |
| Azerbaijan | 17.2 | 15.9 | 1.3 | 8.9 | 8.9 | - | 8.3 | 7.0 | 1.3 |
| Bahrain | 4.2 | 2.8 | 1.3 | 2.6 | 2.3 | 0.3 | 1.5 | 0.5 | 1.0 |
| Bangladesh | 222.6 | 222.6 | - | 52.9 | 52.9 | - | 169.6 | 169.6 | - |
| Barbados | 0.5 | 0.5 | 0.0 | 0.3 | 0.3 | - | 0.3 | 0.2 | 0.0 |
| Belarus | 14.9 | 14.9 | - | 4.1 | 4.1 | - | 10.7 | 10.7 | - |
| Belgium | 23.3 | 19.8 | 3.5 | 16.9 | 16.7 | 0.2 | 6.4 | 3.1 | 3.3 |
| Belize | 0.6 | 0.6 | - | 0.3 | 0.3 | - | 0.3 | 0.3 | - |
| Benin | 12.5 | 12.5 | - | 0.2 | 0.2 | - | 12.2 | 12.2 | - |
| Bhutan | 1.2 | 1.2 | - | 1.1 | 1.1 | - | 0.1 | 0.1 | - |
| Bolivia | 15.3 | 15.3 | - | 6.9 | 6.9 | - | 8.4 | 8.4 | - |
| Bosnia and Herzegovina | 5.8 | 5.8 | - | 1.2 | 1.2 | - | 4.6 | 4.6 | - |
| Botswana | 2.9 | 2.9 | - | 0.7 | 0.7 | - | 2.2 | 2.2 | - |
| Brazil | 382.7 | 349.3 | 33.4 | 245.1 | 243.0 | 2.1 | 137.6 | 106.3 | 31.3 |
| Brunei | 0.7 | 0.7 | 0.0 | 0.5 | 0.5 | - | 0.2 | 0.2 | 0.0 |
| Bulgaria | 14.1 | 12.3 | 1.8 | 2.6 | 2.6 | - | 11.6 | 9.7 | 1.8 |
| Burkina Faso | 20.4 | 20.4 | - | 0.3 | 0.3 | - | 20.1 | 20.1 | - |
| Burundi | 3.5 | 3.5 | - | 0.0 | 0.0 | - | 3.5 | 3.5 | - |
| Cambodia | 39.8 | 29.2 | 10.7 | 24.4 | 23.5 | 0.9 | 15.4 | 5.6 | 9.8 |

| | | | | | | | | | |
|---|---|---|---|---|---|---|---|---|---|
| Cameroon | 27.2 | 27.2 | - | 0.5 | 0.5 | - | 26.7 | 26.7 | - |
| Canada | 66.9 | 65.2 | 1.7 | 56.8 | 56.8 | - | 10.0 | 8.3 | 1.7 |
| Cabo Verde | 0.7 | 0.7 | - | 0.4 | 0.4 | - | 0.3 | 0.3 | - |
| Central African Republic | 1.5 | 1.5 | - | 0.2 | 0.2 | - | 1.3 | 1.3 | - |
| Chad | 20.1 | 20.1 | - | 0.2 | 0.2 | - | 19.9 | 19.9 | - |
| Chile | 45.4 | 34.9 | 10.5 | 33.2 | 29.5 | 3.7 | 12.2 | 5.4 | 6.7 |
| China | 2713.2 | 2437.5 | 275.7 | 2217.6 | 2217.6 | - | 495.6 | 220.0 | 275.7 |
| Colombia | 94.4 | 82.4 | 12.0 | 42.3 | 42.3 | 0.0 | 52.1 | 40.1 | 12.0 |
| Comoros | 1.0 | 1.0 | - | 0.4 | 0.4 | - | 0.6 | 0.6 | - |
| Congo | 5.8 | 5.8 | - | 0.4 | 0.4 | - | 5.4 | 5.4 | - |
| Cook Islands | 0.0 | 0.0 | - | 0.0 | 0.0 | - | 0.0 | 0.0 | - |
| Costa Rica | 8.5 | 8.5 | - | 5.8 | 5.8 | - | 2.7 | 2.7 | - |
| Cote d'Ivoire | 27.3 | 27.3 | - | 2.5 | 2.5 | - | 24.8 | 24.8 | - |
| Croatia | 6.9 | 6.9 | - | 3.5 | 3.5 | - | 3.4 | 3.4 | - |
| Cuba | 22.2 | 22.2 | - | 22.3 | 16.7 | 5.7 | 5.6 | 5.6 | - |
| Cyprus | 2.5 | 2.1 | 0.4 | 1.2 | 1.2 | 0.0 | 1.3 | 0.9 | 0.4 |
| Czechia | 28.1 | 18.5 | 9.6 | 11.9 | 11.8 | 0.0 | 16.3 | 6.7 | 9.6 |
| Democratic Republic of the Congo | 31.0 | 31.0 | - | 0.1 | 0.1 | - | 30.9 | 30.9 | - |

| | | | | | | | | | |
|---|---|---|---|---|---|---|---|---|---|
| Denmark | 11.6 | 10.0 | 1.6 | 8.8 | 8.7 | 0.1 | 2.7 | 1.2 | 1.5 |
| Djibouti | 1.3 | 1.3 | - | 0.1 | 0.1 | - | 1.2 | 1.2 | - |
| Dominica | 0.1 | 0.1 | - | 0.0 | 0.0 | - | 0.1 | 0.1 | - |
| Dominican Republic | 24.9 | 16.5 | 8.5 | 12.1 | 11.1 | 1.0 | 12.9 | 5.4 | 7.5 |
| Ecuador | 27.6 | 27.5 | 0.1 | 21.2 | 21.2 | - | 6.4 | 6.3 | 0.1 |
| Egypt | 124.7 | 124.7 | - | 19.8 | 19.8 | - | 104.9 | 104.9 | - |
| El Salvador | 15.8 | 11.4 | 4.4 | 7.9 | 7.7 | 0.2 | 7.9 | 3.6 | 4.2 |
| Equatorial Guinea | 1.6 | 1.6 | - | 0.4 | 0.4 | - | 1.2 | 1.2 | - |
| Eritrea | - | - | - | - | - | - | - | - | - |
| Estonia | 3.2 | 2.1 | 1.1 | 1.5 | 1.5 | - | 1.7 | 0.6 | 1.1 |
| Eswatini | 1.6 | 1.6 | - | - | - | - | - | - | - |
| Ethiopia | 54.3 | 54.3 | - | 3.9 | 3.9 | - | 50.4 | 50.4 | - |
| Fiji | 1.2 | 1.2 | - | 1.1 | 1.1 | - | 0.2 | 0.2 | - |
| Finland | 9.9 | 9.5 | 0.4 | 7.8 | 7.8 | - | 2.1 | 1.7 | 0.4 |
| France | 98.1 | 77.8 | 20.3 | 96.0 | 94.5 | 1.5 | 18.9 | 0.0 | 18.9 |
| Gabon | 3.1 | 3.1 | - | 0.2 | 0.2 | - | 2.9 | 2.9 | - |
| Georgia | 6.0 | 6.0 | - | 1.8 | 1.8 | - | 4.1 | 4.1 | - |
| Germany | 159.0 | 142.9 | 16.1 | 108.8 | 107.9 | 1.0 | 50.1 | 35.0 | 15.1 |
| Ghana | 35.2 | 35.2 | - | 2.2 | 2.2 | - | 33.0 | 33.0 | - |
| Greece | 21.8 | 18.7 | 3.1 | 12.3 | 12.3 | - | 9.5 | 6.4 | 3.1 |

| | | | | | | | | | |
|---|---|---|---|---|---|---|---|---|---|
| Grenada | 0.2 | 0.2 | - | 0.1 | 0.1 | - | 0.1 | 0.1 | - |
| Guatemala | 20.7 | 20.7 | - | 7.5 | 7.5 | - | 13.2 | 13.2 | - |
| Guinea | 13.1 | 13.1 | - | 1.9 | 1.9 | - | 11.2 | 11.2 | - |
| Guinea-Bissau | 2.0 | 2.0 | - | 0.1 | 0.1 | - | 1.9 | 1.9 | - |
| Guyana | 1.0 | 1.0 | - | 0.6 | 0.6 | - | 0.5 | 0.5 | - |
| Haiti | 14.0 | 14.0 | - | 0.1 | 0.1 | - | 13.9 | 13.9 | - |
| Honduras | 13.3 | 13.3 | - | 5.9 | 5.9 | - | 7.4 | 7.4 | - |
| Hungary | 24.9 | 16.9 | 8.0 | 12.5 | 11.6 | 0.9 | 12.5 | 5.4 | 7.1 |
| Iceland | 0.7 | 0.6 | 0.1 | 0.6 | 0.5 | 0.1 | 0.1 | 0.1 | 0.0 |
| India | 1836.9 | 1836.9 | - | 927.2 | 927.2 | - | 909.7 | 909.7 | - |
| Indonesia | 423.4 | 422.7 | 0.8 | 151.5 | 151.5 | - | 272.0 | 271.2 | 0.8 |
| Iran | 119.7 | 119.7 | - | 61.5 | 61.5 | - | 58.2 | 58.2 | - |
| Iraq | 45.0 | 45.0 | - | 8.1 | 8.1 | - | 36.9 | 36.9 | - |
| Ireland | 8.8 | 8.1 | 0.7 | 7.5 | 7.5 | - | 1.3 | 0.6 | 0.7 |
| Israel | 19.7 | 13.0 | 6.7 | 15.6 | 11.9 | 3.7 | 4.1 | 1.1 | 3.0 |
| Italy | 133.5 | 107.8 | 25.7 | 85.8 | 85.7 | 0.1 | 47.7 | 22.1 | 25.6 |
| Jamaica | 4.4 | 4.4 | - | 0.8 | 0.8 | - | 3.6 | 3.6 | - |
| Japan | 226.3 | 226.3 | - | 172.1 | 172.1 | - | 54.2 | 54.2 | - |
| Jordan | 14.6 | 14.6 | - | 7.1 | 7.1 | - | 7.5 | 7.5 | - |
| Kazakhstan | 42.6 | 28.4 | 14.2 | 14.6 | 14.6 | - | 28.1 | 13.9 | 14.2 |
| Kenya | 58.6 | 58.6 | - | 4.0 | 4.0 | - | 54.6 | 54.6 | - |

| | | | | | | | | | |
|---|---|---|---|---|---|---|---|---|---|
| Kiribati | 0.1 | 0.1 | - | 0.0 | 0.0 | - | 0.1 | 0.1 | - |
| Kuwait | 10.3 | 7.1 | 3.2 | - | - | - | 3.2 | - | 3.2 |
| Kyrgyzstan | 7.8 | 7.8 | - | 1.5 | 1.5 | - | 6.3 | 6.3 | - |
| Laos | 9.0 | 9.0 | - | 5.2 | 5.2 | - | 3.8 | 3.8 | - |
| Latvia | 3.5 | 3.1 | 0.5 | 1.6 | 1.6 | - | 1.9 | 1.5 | 0.5 |
| Lebanon | 10.1 | 9.8 | 0.3 | 3.0 | 3.0 | - | 7.1 | 6.8 | 0.3 |
| Lesotho | 2.8 | 2.8 | - | 0.4 | 0.4 | - | 2.4 | 2.4 | - |
| Liberia | 5.3 | 5.3 | - | 0.1 | 0.1 | - | 5.3 | 5.3 | - |
| Libya | 9.0 | 9.0 | - | 1.7 | 1.7 | - | 7.3 | 7.3 | - |
| Lithuania | 5.8 | 4.7 | 1.1 | 3.3 | 3.2 | 0.0 | 2.6 | 1.5 | 1.0 |
| Luxembourg | 1.2 | 1.1 | 0.1 | 0.8 | 0.8 | 0.0 | 0.4 | 0.3 | 0.1 |
| Macedonia | 4.1 | 3.6 | 0.5 | 1.5 | 1.5 | 0.0 | 2.5 | 2.1 | 0.5 |
| Madagascar | 29.5 | 29.5 | - | 0.4 | 0.4 | - | 29.1 | 29.1 | - |
| Malawi | 19.1 | 19.1 | - | 1.1 | 1.1 | - | 18.0 | 18.0 | - |
| Malaysia | 74.1 | 50.9 | 23.2 | 46.1 | 46.1 | 0.0 | 28.0 | 4.8 | 23.2 |
| Maldives | 0.9 | 0.9 | - | 0.7 | 0.7 | - | 0.2 | 0.2 | - |
| Mali | 18.6 | 18.6 | - | 0.5 | 0.5 | - | 18.2 | 18.2 | - |
| Malta | 0.9 | 0.8 | 0.1 | 0.8 | 0.8 | 0.0 | 0.1 | 0.0 | 0.1 |
| Marshall Islands | 0.1 | 0.1 | - | 0.0 | 0.0 | 0.0 | 0.0 | 0.0 | - |
| Mauritania | 6.2 | 6.2 | - | 1.2 | 1.2 | - | 5.0 | 5.0 | - |
| Mauritius | 2.2 | 2.2 | - | 1.7 | 1.7 | - | 0.5 | 0.5 | - |

| | | | | | | | | | |
|---|---|---|---|---|---|---|---|---|---|
| Mexico | 144.9 | 144.9 | - | 106.1 | 106.1 | - | 38.8 | 38.8 | - |
| Micronesia | 0.2 | 0.2 | 0.0 | 0.1 | 0.1 | - | 0.1 | 0.1 | 0.0 |
| Moldova | 6.5 | 6.5 | - | 1.4 | 1.4 | - | 5.1 | 5.1 | - |
| Monaco | 0.1 | 0.1 | - | - | - | - | - | - | - |
| Mongolia | 4.8 | 4.8 | - | 4.4 | 4.4 | - | 0.5 | 0.5 | - |
| Montenegro | 1.2 | 1.1 | 0.2 | 0.5 | 0.5 | - | 0.8 | 0.6 | 0.2 |
| Morocco | 56.3 | 56.3 | - | 42.8 | 42.8 | - | 13.5 | 13.5 | - |
| Mozambique | 32.6 | 32.6 | - | 3.7 | 3.7 | - | 28.9 | 28.9 | - |
| Myanmar | 15.9 | 15.9 | - | 12.3 | 12.3 | - | 3.7 | 3.7 | - |
| Namibia | 2.9 | 2.9 | - | 0.5 | 0.5 | - | 2.4 | 2.4 | - |
| Nauru | 0.0 | 0.0 | - | - | - | - | - | - | - |
| Nepal | 37.7 | 37.7 | - | 14.4 | 14.4 | - | 23.3 | 23.3 | - |
| Netherlands | 28.7 | 28.0 | 0.7 | 23.9 | 23.9 | - | 4.8 | 4.1 | 0.7 |
| New Zealand | 8.2 | 8.0 | 0.2 | 5.6 | 5.6 | - | 2.6 | 2.4 | 0.2 |
| Nicaragua | 8.6 | 8.6 | - | 1.0 | 1.0 | 0.0 | 7.6 | 7.6 | - |
| Niger | 21.1 | 21.1 | - | 0.6 | 0.6 | - | 20.4 | 20.4 | - |
| Nigeria | 208.0 | 208.0 | - | 7.1 | 7.1 | - | 200.9 | 200.9 | - |
| Niue | 0.0 | 0.0 | - | - | - | - | - | - | - |
| North Korea | - | - | - | - | - | - | - | - | - |
| Norway | 10.4 | 9.4 | 1.0 | 7.9 | 7.9 | - | 2.5 | 1.5 | 1.0 |
| Oman | 8.3 | 8.3 | - | 5.2 | 5.2 | - | 3.1 | 3.1 | - |
| Pakistan | 303.2 | 303.2 | - | 89.7 | 89.7 | - | 213.5 | 213.5 | - |

| | | | | | | | | | |
|---|---|---|---|---|---|---|---|---|---|
| Palau | 0.0 | 0.0 | 0.0 | - | - | - | 0.0 | - | 0.0 |
| Panama | 8.2 | 6.8 | 1.4 | 5.2 | 5.2 | - | 2.9 | 1.5 | 1.4 |
| Papua New Guinea | 10.5 | 10.5 | - | 0.2 | 0.2 | - | 10.3 | 10.3 | - |
| Paraguay | 9.4 | 9.4 | - | 4.8 | 4.8 | 0.0 | 4.5 | 4.5 | - |
| Peru | 46.9 | 46.7 | 0.1 | 29.9 | 29.9 | - | 17.0 | 16.8 | 0.1 |
| Philippines | 161.8 | 161.8 | - | 48.9 | 48.9 | - | 112.8 | 112.8 | - |
| Poland | 79.8 | 64.4 | 15.4 | 37.8 | 37.8 | - | 42.0 | 26.6 | 15.4 |
| Portugal | 19.7 | 17.1 | 2.7 | 16.1 | 16.1 | - | 3.7 | 1.0 | 2.7 |
| Qatar | 5.9 | 5.1 | 0.8 | 4.7 | 4.7 | - | 1.1 | 0.3 | 0.8 |
| Romania | 50.7 | 33.8 | 16.9 | 10.6 | 10.6 | - | 40.1 | 23.2 | 16.9 |
| Russia | 340.9 | 226.2 | 114.8 | 93.9 | 93.0 | 0.9 | 247.0 | 133.1 | 113.9 |
| Rwanda | 13.2 | 13.2 | - | 3.8 | 3.8 | - | 9.3 | 9.3 | - |
| Saint Kitts and Nevis | 0.1 | 0.1 | - | 0.0 | 0.0 | 0.0 | 0.0 | 0.0 | - |
| Saint Lucia | 0.3 | 0.3 | - | 0.1 | 0.1 | - | 0.2 | 0.2 | - |
| Saint Vincent and the Grenadines | 0.2 | 0.2 | - | 0.0 | 0.0 | - | 0.1 | 0.1 | - |
| Samoa | 0.2 | 0.2 | - | 0.2 | 0.2 | - | 0.0 | 0.0 | - |
| San Marino | 0.0 | 0.0 | - | 0.0 | 0.0 | - | 0.0 | 0.0 | - |
| Sao Tome and | 0.2 | 0.2 | - | 0.1 | 0.1 | - | 0.1 | 0.1 | - |

| | | | | | | | | | |
|---|---|---|---|---|---|---|---|---|---|
| Principe | | | | | | | | | |
| Saudi Arabia | 55.2 | 53.2 | 2.0 | 43.1 | 43.1 | - | 12.1 | 10.1 | 2.0 |
| Senegal | 17.0 | 17.0 | - | 1.8 | 1.8 | - | 15.2 | 15.2 | - |
| Serbia | 22.5 | 15.4 | 7.1 | 6.7 | 6.0 | 0.7 | 15.8 | 9.4 | 6.4 |
| Seychelles | 0.2 | 0.2 | - | 0.2 | 0.2 | - | 0.0 | 0.0 | - |
| Sierra Leone | 8.4 | 8.4 | - | 0.3 | 0.3 | - | 8.1 | 8.1 | - |
| Singapore | 10.3 | 10.3 | - | 9.6 | 9.2 | 0.4 | 1.1 | 1.1 | - |
| Slovakia | 14.2 | 9.4 | 4.8 | 4.7 | 4.7 | - | 9.5 | 4.7 | 4.8 |
| Slovenia | 3.2 | 2.5 | 0.7 | 2.2 | 2.2 | - | 1.0 | 0.4 | 0.7 |
| Solomon Islands | 0.7 | 0.7 | - | 0.1 | 0.1 | - | 0.6 | 0.6 | - |
| Somalia | 14.9 | 14.9 | - | 0.5 | 0.5 | - | 14.3 | 14.3 | - |
| South Africa | 93.4 | 90.9 | 2.5 | 18.6 | 18.6 | - | 74.8 | 72.3 | 2.5 |
| South Korea | 89.6 | 86.2 | 3.4 | 67.7 | 67.7 | 0.0 | 22.0 | 18.5 | 3.4 |
| South Sudan | 11.7 | 11.7 | - | 0.1 | 0.1 | - | 11.5 | 11.5 | - |
| Spain | 86.4 | 77.3 | 9.0 | 70.7 | 70.4 | 0.3 | 15.7 | 6.9 | 8.7 |
| Sri Lanka | 36.0 | 34.1 | 1.9 | 26.7 | 26.7 | - | 9.3 | 7.4 | 1.9 |
| Sudan | 46.9 | 46.9 | - | 1.5 | 1.5 | - | 45.4 | 45.4 | - |
| Suriname | 0.8 | 0.8 | - | 0.4 | 0.4 | - | 0.4 | 0.4 | - |
| Sweden | 18.1 | 17.1 | 1.0 | 13.9 | 13.9 | - | 4.2 | 3.2 | 1.0 |
| Switzerland | 15.3 | 15.0 | 0.3 | 10.6 | 10.6 | - | 4.7 | 4.4 | 0.3 |
| Syria | 22.3 | 22.3 | - | 0.9 | 0.9 | - | 21.5 | 21.5 | - |

| | | | | | | | | | |
|---|---|---|---|---|---|---|---|---|---|
| Tajikistan | 11.0 | 11.0 | - | 4.5 | 4.5 | - | 6.5 | 6.5 | - |
| Tanzania | 59.4 | 59.4 | - | 0.9 | 0.9 | - | 58.5 | 58.5 | - |
| Thailand | 182.2 | 121.5 | 60.7 | 57.4 | 55.8 | 1.6 | 124.9 | 65.7 | 59.1 |
| The Bahamas | 0.7 | 0.7 | - | 0.2 | 0.2 | - | 0.4 | 0.4 | - |
| The Gambia | 0.7 | 0.7 | - | 0.2 | 0.2 | - | 0.5 | 0.5 | - |
| Timor-Leste | 1.5 | 1.5 | - | 0.8 | 0.8 | - | 0.7 | 0.7 | - |
| Togo | 8.0 | 8.0 | - | 1.2 | 1.2 | - | 6.8 | 6.8 | - |
| Tonga | 0.1 | 0.1 | - | 0.1 | 0.1 | - | 0.0 | 0.0 | - |
| Trinidad and Tobago | 2.3 | 2.3 | - | 1.1 | 1.1 | - | 1.2 | 1.2 | - |
| Tunisia | 20.9 | 17.0 | 3.9 | 8.3 | 8.3 | - | 12.5 | 8.6 | 3.9 |
| Turkey | 154.0 | 133.7 | 20.3 | 111.7 | 100.2 | 11.5 | 42.3 | 33.6 | 8.8 |
| Turkmenistan | 7.8 | 7.8 | - | - | - | - | - | - | - |
| Tuvalu | 0.0 | 0.0 | - | - | - | - | - | - | - |
| Uganda | 56.6 | 56.6 | - | 2.6 | 2.6 | - | 54.0 | 54.0 | - |
| Ukraine | 75.5 | 75.5 | - | 13.4 | 13.4 | - | 62.1 | 62.1 | - |
| United Arab Emirates | 21.2 | 19.0 | 2.2 | 20.4 | 20.4 | - | 2.2 | 0.0 | 2.2 |
| United Kingdom | 147.3 | 115.0 | 32.3 | 94.2 | 94.2 | - | 53.1 | 20.7 | 32.3 |
| United States | 660.0 | 558.2 | 101.8 | 399.6 | 392.8 | 6.8 | 260.5 | 165.4 | 95.0 |
| Uruguay | 10.0 | 5.7 | 4.3 | 6.4 | 5.3 | 1.0 | 3.7 | 0.4 | 3.3 |

| | | | | | | | | | |
|---|---|---|---|---|---|---|---|---|---|
| Uzbekistan | 51.1 | 51.1 | - | 22.0 | 22.0 | - | 29.1 | 29.1 | - |
| Vanuatu | 0.3 | 0.3 | - | 0.1 | 0.1 | - | 0.3 | 0.3 | - |
| Venezuela | 38.3 | 38.3 | - | 16.1 | 16.1 | - | 22.2 | 22.2 | - |
| Vietnam | 157.9 | 157.9 | - | 50.6 | 50.6 | - | 107.3 | 107.3 | - |
| Yemen | 32.6 | 32.6 | - | 0.4 | 0.4 | - | 32.2 | 32.2 | - |
| Zambia | 18.0 | 18.0 | - | 0.8 | 0.8 | - | 17.2 | 17.2 | - |
| Zimbabwe | 18.0 | 18.0 | - | 5.5 | 5.5 | - | 12.5 | 12.5 | - |

Abbreviation: AFR, African Region; AMR, Region of Americas; EMR, Eastern Mediterranean Region; EUR, European Region; SEAR, South-East Asia Region; WPR, Western Pacific Region.
